# Genome-wide association study investigating short and long sleep duration: a cross-population meta-analysis

**DOI:** 10.1101/2022.09.09.22279703

**Authors:** Isabelle Austin-Zimmerman, Daniel F. Levey, Olga Giannakopoulou, Joseph D. Deak, Marco Galimberti, Hang Zhou, Spiros Denaxas, Haritz Irizar, Karoline Kuchenbaecker, Andrew McQuillin, the Million Veteran Program, John Concato, Daniel J. Buysse, J. Michael Gaziano, Daniel J. Gottlieb, Renato Polimanti, Murray B. Stein, Elvira Bramon, Joel Gelernter

## Abstract

Sleep duration has been linked to a wide range of negative health outcomes and to reduced life expectancy. We conducted genome-wide association studies of short (≤5 hours) and long (≥10 hours) sleep duration in adults of European, African, East Asian, and admixed-American ancestry from UK Biobank and the Million Veteran Program. In a cross-population meta-analysis we identified 84 independent loci for short sleep and 1 for long sleep. We estimated SNP-based heritability for both sleep traits in each ancestry based on population derived linkage disequilibrium (LD) scores using cov-LDSC. We identified positive genetic correlation between short and long sleep traits (r_g_ = 0.16±0.04; P=0.0002), as well as similar patterns of genetic correlation with other psychiatric and cardiometabolic phenotypes. Mendelian randomisation revealed a directional causal relationship between short sleep and depression, and a bidirectional causal relationship between long sleep and depression.

## Introduction

Sleep is one of the most highly conserved traits across the animal kingdom, indicating a strong evolutionary requirement. It is an essential and fundamental property of neurons and networks across the brain (1,2). Sleep occurs in any organism with even a very simple neuronal/glial network (e.g. *Cassiopeia, C. elegans*), and is preserved in subjects surviving lesions in any brain region (1,3–5). However, many of the molecular processes underlying sleep remain unclear.

Human sleep can be characterized along dimensions such as duration, timing, efficiency, and regularity, each sometimes associated with adverse health outcomes. However, sleep duration has been most widely studied, and relates to outcomes including obesity, cardiovascular disease, and mortality (6). Both unusually long and unusually short sleep duration have been related to multiple psychiatric conditions, including major depressive disorder (MDD), anxiety, and psychosis, though the causal relationship between sleep duration and these disorders is not established (7–10).

Genetic research, and in particular genome-wide association studies (GWAS), may help elucidate some of the biological processes that underlie variability in sleep across individuals, by identifying risk loci associated with higher or lower than average sleep duration. Self-reported sleep duration is a complex trait, with a genetic component established through twin and family studies as well as several GWAS (8,9,11–14). A recent GWAS in 446,118 European-ancestry (EUR) UK Biobank participants identified over 70 independent genetic loci associated with habitual, self-reported sleep duration (measured as a continuous trait reported in hour increments), as well as several linked specifically to unusually long (nine hours or more) and short (six hours or less) sleep duration (7). SNP-based heritability of sleep duration was reported to be 9.8%. This study, and several others, identified common variants at or near the *VRK2* and *PAX8* genes (8,15,16). *VRK2* encodes a serine/threonine kinase protein which is essential to multiple signal transduction pathways (7,14,17). Single nucleotide polymorphisms (SNPs) within this gene have been associated with a range of psychiatric disorders, such as schizophrenia and depression, as well as epilepsy and some cardiometabolic traits. *PAX8* is a transcription factor important in the development and function of the thyroid.

Like many GWAS, these studies have been conducted primarily in EUR participants. Replicating these findings in other populations, or identifying ancestry-specific risk loci, is essential for furthering our understanding of the biological mechanisms behind sleep, and the effects of sleep on biology.

The UK Biobank and Million Veteran Program (MVP) represent two of the world’s largest biobanks, both containing genetic data and a wide range of environmental and medical information. UK Biobank is a population-based study, including over 500,000 UK-based participants (18,19). MVP is a US military sample, having recruited so far over 825,000 veterans (20). We conducted a cross-population meta-analysis of short and long sleep duration using GWAS results from UK Biobank and MVP. This study aimed to build on the existing understanding of the genetics of sleep duration and to take advantage of the diverse populations included in UK Biobank and MVP to consider risk loci across multiple populations, as well as ancestry-specific regions of interest.

## Methods

### Participants

The UK Biobank and MVP cohorts have been described previously (18–20). The UK Biobank study was approved by the North-West Research Ethics Committee (ref 06/MREC08/65) in accordance with the Declaration of Helsinki. Research involving the MVP in general is approved by the VA Central Institutional Review Board. All participants in both cohorts provided written informed consent.

### Genotyping, imputation, and quality control

#### UK Biobank

Genotyping and imputation of UK Biobank subjects has been described previously (18). Briefly, genotyping for UK Biobank participants was undertaken using the Affymetrix UK BiLEVE Axiom array (used for the first ∼50,000 participants) and the Affymetrix UK Biobank Axiom Array (∼450,000 participants) (21). These arrays are >95% similar and include ∼820,000 SNP and indel markers (http://www.ukbiobank.ac.uk/). Quality control and imputation of over 90 million SNPs, indels and large structural variants were performed centrally (21). Samples identified as outliers for heterozygosity and/or missingness were removed, leaving a total sample of 487,411. The fully imputed genetic data used in this study, with basic sample and variant level quality control as reported in reference (21), were made available in March 2018.

Additional local post-imputation SNP-level quality control was conducted to remove SNPs with a imputation INFO score <0.3 or those with minor allele frequency (MAF) <0.01. Further individual-level quality control was conducted locally to remove samples with mismatch between reported sex and genetically inferred sex (due to risk of sample processing errors) and those with excessive genetic relatedness (greater than 10 third-degree relatives based on kinship calculations provided centrally by UK Biobank). Individuals missing either sleep or essential quality data were excluded. The final list was then checked to remove those who had withdrawn consent.

Genetic ancestry of the UK Biobank sample was assessed using principal component analysis (PCA) in combination with self-reported ethnicity data. A list of 409,728 EUR individuals was identified centrally by the UK Biobank (18). Further local analysis was conducted to delineate the ancestry of another 77,683 participants from diverse populations, applying the same thresholds as described in (18). Two rounds of PCA were performed using the PC-AiR algorithm (22), that captures population structure. Relatedness in this sample was assessed using PC-Relate and the ancestry representative PCs (23). Of the samples that passed the QC procedures described here, over 99% provided self-reported sleep duration data.

#### MVP

Genotyping and imputation of MVP participants has been described previously (20). Briefly, MVP subjects were genotyped using a customised Affymetrix Axiom Array, similar to the UK Biobank array. MVP genotype data was imputed using Minimac4 and a reference panel from the African Genome Resources (AGR) panel by the Sanger Institute. Indels and complex variants were imputed independently from the 1000 Genomes phase 3 panel (G1K) and merged in a similar approach to UKB HRC + UK10K. SNPs with an imputation info score <0.3, estimated genotype hard call missingness rate of >0.2 or a MAF < 0.001 were removed. PCA was conducted using Eigensoft (24). Genetic ancestry of the MVP participants was assigned separately within each data tranche, based upon the first 10 principal components, with 1000 Genomes Project (phase 3) EUR, African (AFR), Admixed American (AMR) and East Asian (EAS) data as reference samples (20).

### Phenotypic assessment and covariate measures

In both cohorts, we selected participants who had provided self-reported data on sleep duration. In UK Biobank, participants were asked “About how many hours sleep do you get in every 24h? (Please include naps)” as part of the baseline assessment. Responses were given in hour increments and participants who claimed to sleep less than three hours or more than 12 were prompted to confirm their answer. Data on sleep duration was collected from the MVP lifestyle questionnaire, where participants were asked “How many hours do you usually sleep each day (24-hour period)?”. The response options were multiple choice: 5 or less, 6, 7, 8, 9 or 10 or more.

Due to this difference in how the sleep duration data were recorded, we could not consider sleep as a continuous variable in both samples. In addition, we did not wish to assume that short and long sleep are necessarily on the same biological continuum. We therefore defined ‘short’ sleep duration as those reporting ≤5 hours sleep, ‘normal’ as 7-8 hours and ‘long’ as ≥10 hours (25).

In both UK Biobank and MVP samples we included sex, age at recruitment, and the first 10 principal components as covariates in the GWAS. In the UK Biobank sample, the genotyping array used was included as an additional covariate. We found that individuals who reported being diagnosed with obstructive sleep apnoea were significantly over-represented in both the short and long sleep groups, and we therefore excluded these participants from our analysis in both the UK Biobank and MVP cohorts.

Although the ‘normal’ or medium sleepers (7-8 hours) represents the largest group in both samples and the distribution of responses is similar, the MVP sample has a significantly greater proportion of both short and long sleepers (Figure 1). UK Biobank is a population-based study, and differences between the sample and the UK population have been described in (26); this could also relate to how the phenotype was elicited. MVP recruits through the US Department of Veterans Affairs (VA) Healthcare System, meaning these participants can be considered a patient population with a broad range of potential health conditions, some of which can be expected to impact sleep duration.

**Figure 1.**
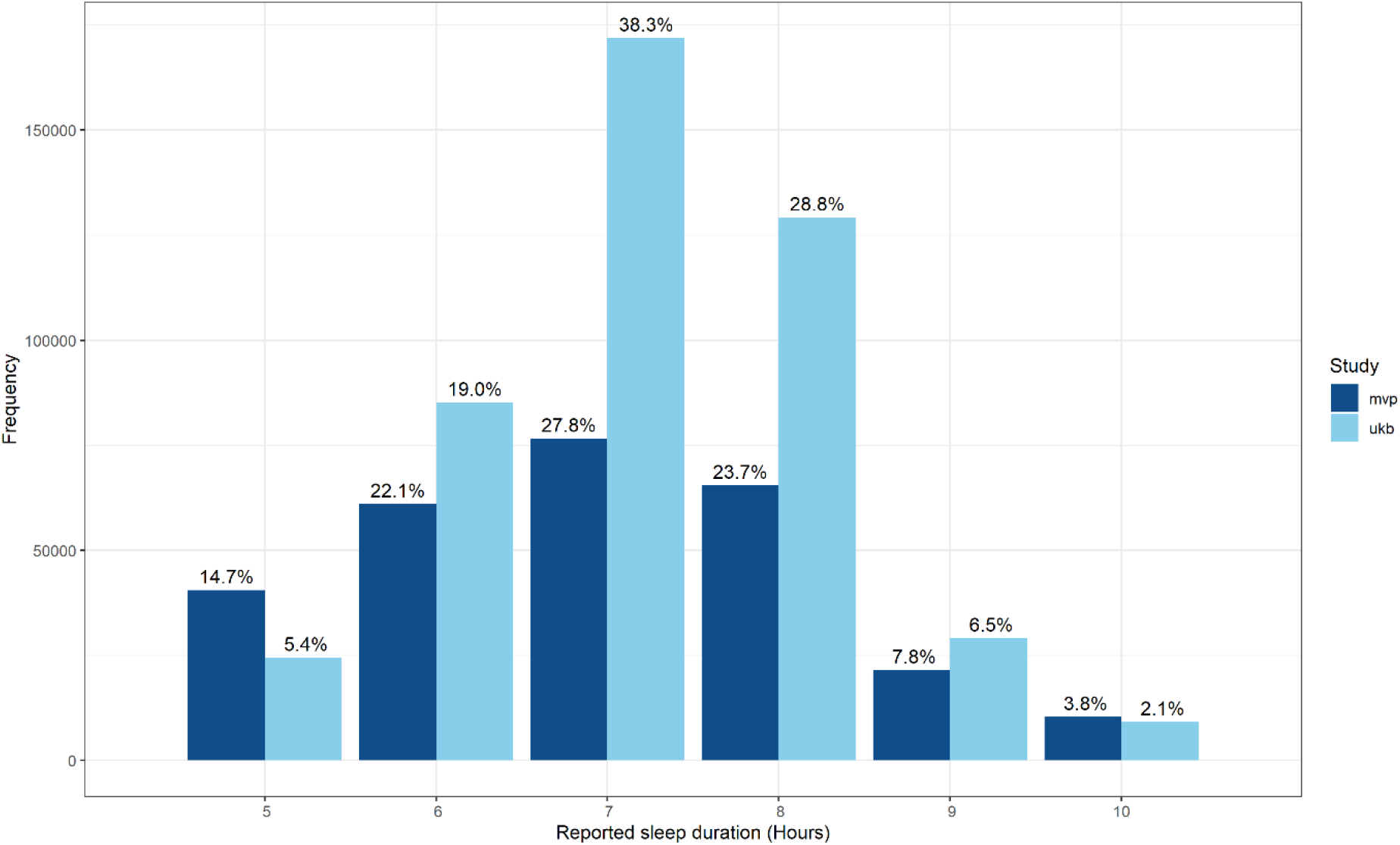
Self-reported sleep duration in UK Biobank and MVP samples

### Hours of daylight exposure

We evaluated possible effects of hours of daylight on sleep hours, as this is known to have a significant impact on reported sleep duration (27–30). We calculated estimated solar irradiance for each participant based on the location of their recruitment site. We downloaded monthly direct normal solar irradiation data from the European Commission Photovoltaic Geographical Information System (31) and the National Solar Radiation Database (32). For UK Biobank subjects, solar irradiation indices were based on recruitment site and the month of their recruitment. For MVP subjects, only average annual data were available. We conducted a linear regression analysis to examine the effects of solar irradiance on sleep hours, including age and sex as covariates. This analysis was conducted using R version 3.5.0 (2018-04-23) (33).

### Statistical analyses

We conducted GWAS on two separate phenotypes, short sleep duration vs normal and long sleep duration vs normal, in each of the independent primary samples (UKB-EUR, UKB-AFR, UKB-EAS, UKB-AMR, MVP-EUR, MVP-AFR, MVP-AMR. GWAS analysis was conducted by logistic regression using PLINK 2.0 on genotype dosage data, including age, sex, first 10 principal components as covariates, and in the case of UK Biobank the genotype array as covariates (34). Where kinship scores showed a relatedness between a pair as closer than 2^rd^ degree, one of each the pair was excluded at random.

We used METAL (35) to conduct independent fixed effect meta-analyses in ancestry specific samples and to conduct a cross-population meta-analysis using all primary GWAS from UK Biobank and MVP. The resulting summary statistics were filtered to remove any SNP that did not appear across all four population groups, in either UK Biobank or MVP data. For each primary GWAS, meta-analysis and cross-population meta-analysis, we calculated LD intercept to assess genomic inflation due to sample size and polygenicity of trait (36). Manhattan and quantile-quantile (Q-Q) plots were created using the R packages ggplot2 and Hudson (33,37). Independent GWAS signals were identified through clumping of results with an r^2^ of 0.6. A second clumping of the independent SNPs was performed with r^2^ of 0.1 to identify lead SNPs (38).

### Gene-based tests

We uploaded summary statistics from the primary GWAS and the meta-analyses into the FUMA (Functional Mapping and Annotation) GWAS platform version 1.3.6, to examine gene-level associations using MAGMA (Multi-Marker Analysis of GenoMic Annotation)(38). The significance threshold was calculated using a Bonferroni multiple testing correction to account for the specific number of protein coding genes in each gene-based test.

### Cross-population transferability of loci

There is often only limited overlap in genome-wide significant (GWS) risk loci across population groups. Differences in LD structure and allele frequency among population groups makes it difficult to determine if an observed association from a primarily European study is replicated in other populations, as the truly causal variant is often unknown (39). Using the R package LDlinkR (40) we developed ‘credible sets’ of SNPs in EAS and AFR populations that are in high LD (r^2^ > 0.6) with the GWS loci from the European analysis of short and long sleep duration. We then searched the UK Biobank/MVP short and long sleep meta-analyses results for EAS and AFR populations for evidence of association (p<0.05).

### Genetic correlation and SNP-based heritability

We used linkage disequilibrium score regression (LDSC) to estimate SNP-based heritability. For the EUR sample, we used reference LD scores provided by the 1000 Genomes Project (41). The high admixture in the AFR sample means that the reference panel data may be unreliable. We therefore used LD scores calculated from the primary genotype data with principal components included as covariates. For the UK Biobank sample, we used the scores published by the Pan-UKBB group. For the MVP sample we used the equivalent cov-LDSC method (described in (42), https://github.com/immunogenomics/cov-ldsc).

Converting the observed SNP-heritability to the liability-scale estimates presented here required an estimation of the population prevalence of ‘cases’ (i.e., short or long sleepers). We based these estimates on the proportion of short and long sleepers in the meta-analysis of UK Biobank and MVP data. These estimates were weighted based on the sample size of the primary studies. Due to observed differences in the distribution of sleep duration, we calculated these estimates separately in the EUR and AFR populations. A higher proportion of AFR subjects report sleeping six hours or less, and a smaller proportion of AFR subjects report sleeping seven, eight, or nine hours, compared to EUR. In the EUR cohort this resulted in an estimated population prevalence (K) of 0.11 for short and K=0.04 for long sleep. In the AFR cohort this resulted in a K=0.43 for short and K=0.05 for long sleep.

For the EUR cohort we used LDSC to calculate genetic correlation between sleep duration and a range of cognitive, neuropsychiatric, cardiac, and metabolic traits (36,43,44). We used LDHub to assess the genetic correlation of short and long sleep to all available traits. In addition, we used LDSC to assess the genetic correlation to traits based upon GWAS summary statistics downloaded from the Psychiatric Genomics Consortium (PGC) website (https://www.med.unc.edu/pgc/results-and-downloads/) and from the Sleep Disorder Knowledge Portal (http://www.kp4cd.org/dataset_downloads/sleep).

Finally, we assessed the genetic correlation between short and long sleep duration based upon the summary statistics for the meta-analyses as described above, using LDSC for the within-ancestry analyses (36) and Popcorn (version 0.9.6: https://github.com/brielin/Popcorn) for the cross-population analyses (45).

### Mendelian randomisation (MR)

We conducted two-sample MR of both short and long sleep duration with the summary statistics of MDD from the Psychiatric Genomics Consortium depression workgroup (46), using the TwoSampleMR package in R (46,47). The genetic instruments were defined as the independent variants that reached a suggestive significance threshold of p<1×10^−5^. To avoid sample overlap, we used a version of the PGC data that excludes UK Biobank samples. We used the inverse-variance weighted (IVW) method as our primary MR model. We also conducted MR-Egger, weighted median, and weighted mode analyses to test for horizonal pleiotropy and potentially invalid genetic instruments (48,49). MR-robust associated profile score (MR-RAPS) was conducted as a further sensitivity analysis to account for potential weak instrument bias or extreme outliers (50). MR analyses were conducted in EUR samples only.

Calculation of the Egger intercept can identify directional pleiotropy which can bias the inverse variance estimates. Where there is directional pleiotropy, the MR-Egger analysis may provide a more reliable effect estimate. Where the Egger-intercept is non-significant, this demonstrates a lack of directional horizontal pleiotropy and provides confidence in the estimates using the inverse variance method. If we identified a significant Egger intercept, we repeated the analyses using only genome-wide significant SNPs (p<5×10^−8^).

## Results

### Sample

Table 1 outlines the age and sex distribution in each ancestry group across the two cohorts. UK Biobank has a higher proportion of women than men. As a US military veteran sample, men are heavily over-represented in MVP, especially in EUR and somewhat less so in the AFR, EAS and AMR samples. Both cohorts are adult samples, with UK Biobank specifically recruiting adults aged 40-70 years. Though there was no age restriction for recruitment to MVP, median age in MVP is higher at 66 years versus 58 years in UK Biobank. AFR participants make up a higher percentage of the overall sample in MVP than UKB. Figure 1 summarises the distribution of sleep hours in each cohort.

### Hours of daylight exposure

In the UK Biobank samples, higher levels of solar irradiation were significantly associated with shorter reported sleep duration, though the effect size is small (estimate = -4.8×10^−4^±5×10^−5^ hours, p < 2×10^−16^). In the MVP sample, based on annual irradiation data rather than monthly data, increased solar irradiation was not significantly associated with sleep duration (estimate = -4.0×10^−3^±3.0×10^−3^ hours, p=0.157).

### Ancestry-specific meta-analyses

A meta-analysis comparing short (n=47,180) versus normal (n=384,594) sleep duration EUR individuals from both cohorts identified 46 genomic risk loci that reached genome-wide significance (GWS) (supplementary figure 17). Of these, 19 were previously associated with a variety of sleep-related phenotypes (see supplementary tables 1-4).

A meta-analysis comparing long (n=15,995) versus normal sleep duration in the EUR participants from both cohorts identified one genome-wide significant locus on chromosome 2 (rs62158206, OR=0.93±0.01, p=3.6×10^−8^) near the *PAX8* gene (supplementary figure 17).

SNP-based heritability (h^2^) was estimated to be 11.9% (p=2.45×10^−115^) for short sleep, and 7.8% (p=1.61×10^−20^) for long sleep. inflation was within the expected range given the sample sizes and polygenicity of the traits in question, with LD intercept close to one (intercept short sleep=1.017±0.01; intercept long sleep=0.99±0.01) (supplementary tables 5 and 6).

A meta-analysis comparing short (n=11,352) versus normal (n=15,305) sleep duration in AFR participants of both cohorts did not identify any GWS loci, though 42 loci reached a suggestive threshold of p≤1×10^−5^, with the strongest association at rs1412139 on chromosome 1 (OR=1.1±0.02, p=1.9×10^−7^) (supplementary figure 19).

A meta-analysis comparing long (n=1,128) versus normal sleep duration in AFR participants from both cohorts identified one GWS locus, rs148926968 on chromosome 13 (OR=0.43±0.1, p=2.6×10^−8^) (supplementary figure 19). SNP-based inflation was within expected range given the sample sizes and polygenicity of the traits in question.

With LD scores calculated from UK Biobank data, we estimate the SNP-based heritability of short sleep duration in AFR to be 8.8 (intercept=1.00±0.01, p=0.04), and using MVP LD score data, we estimate the SNP-based heritability of short sleep duration in AFR to be 7.5% (intercept=0.99±0.01, p=4.0×10^−3^). SNP-based heritability for long sleep was not significant in this sample using either set of LD scores (Table 2 and supplementary tables 5 and 6).

**Table 1.**
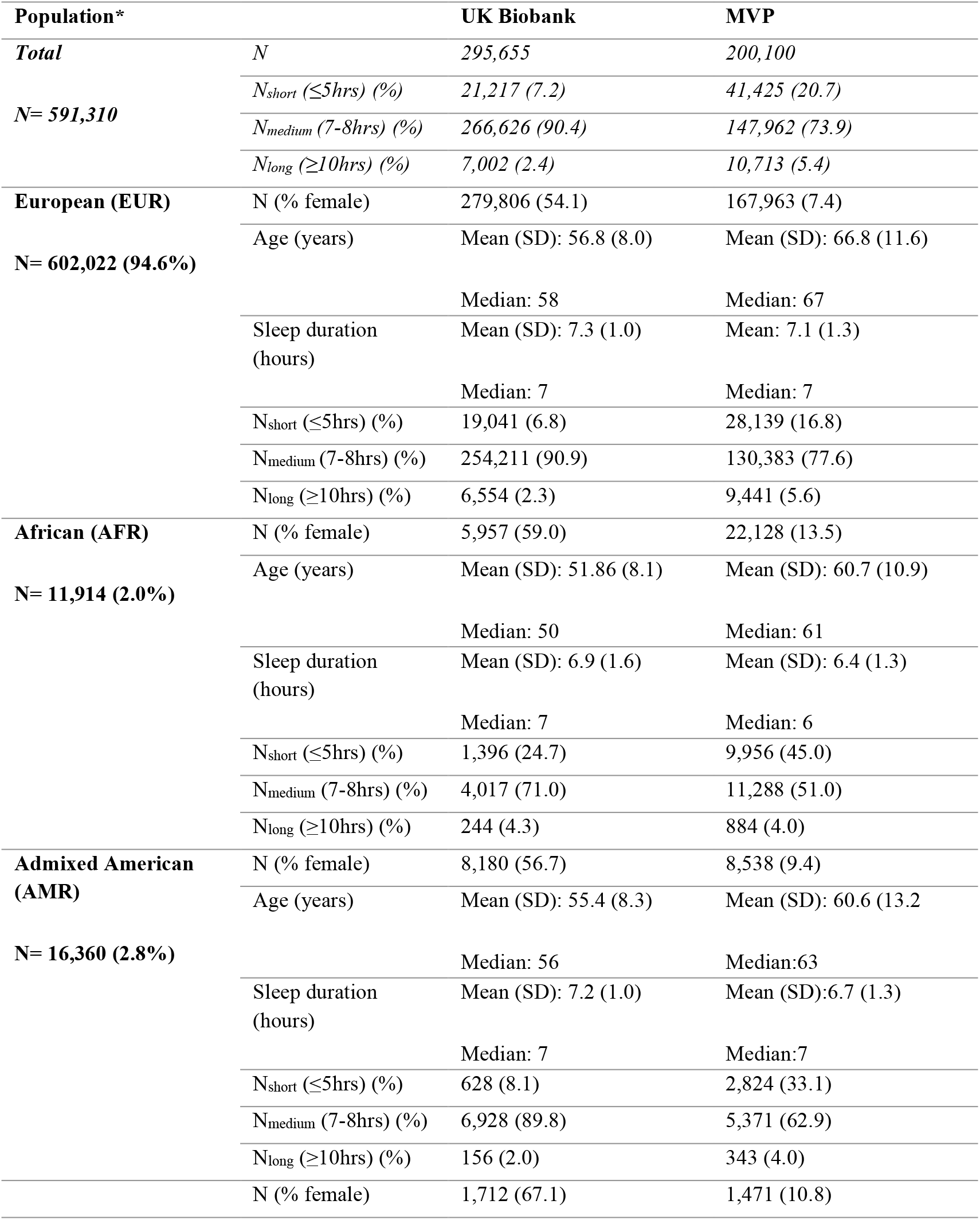
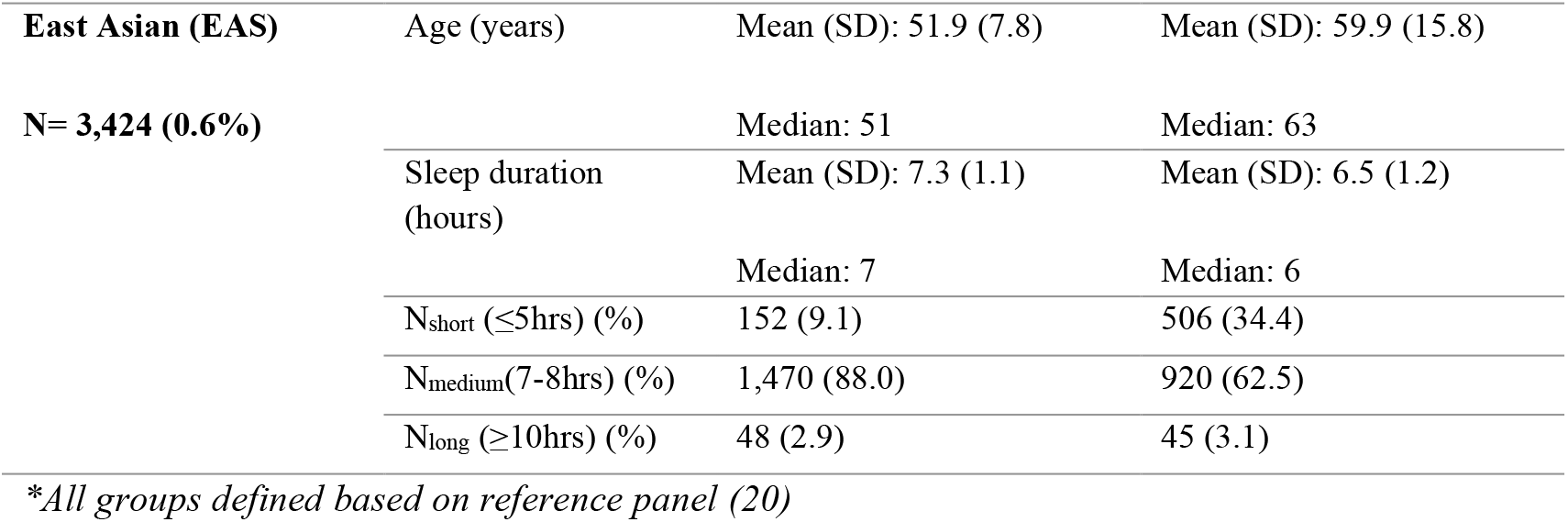
Sample demographics and case/control status

**Table 2.**
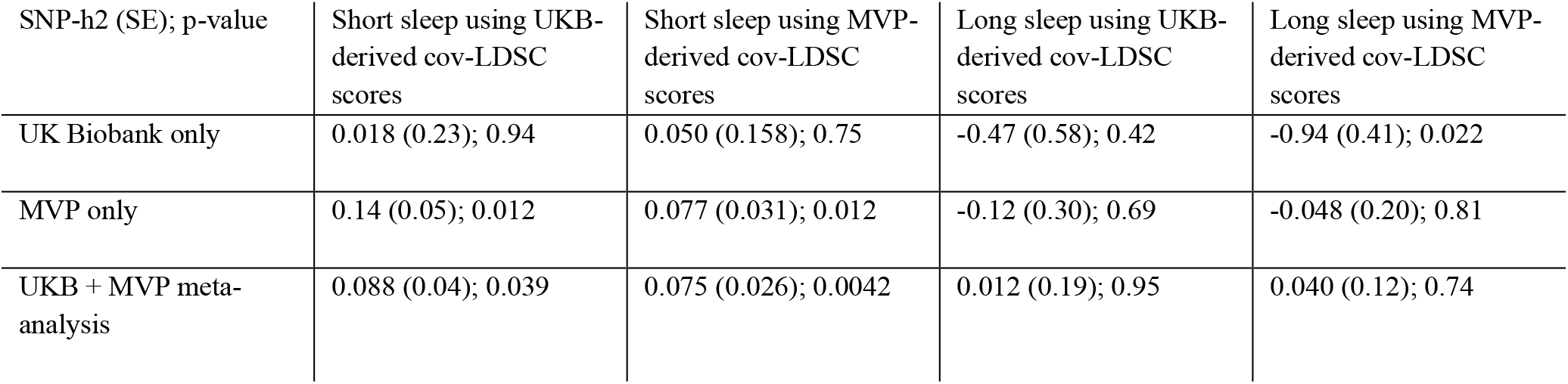
Summary of SNP-heritability estimates (liability scale) for AFR population, derived using LD scores calculated using cov-LDSC, based on either UKB or MVP samples.

| SNP-h<sup>2</sup> (SE); p-value | Short sleep using UKB-derived cov-LDSC scores | Short sleep using MVP-derived cov-LDSC scores | Long sleep using UKB-derived cov-LDSC scores | Long sleep using MVP-derived cov-LDSC scores |
| --- | --- | --- | --- | --- |
| UK Biobank only | 0.018 (0.23); 0.94 | 0.050 (0.158); 0.75 | -0.47 (0.58); 0.42 | -0.94 (0.41); 0.022 |
| MVP only | 0.14 (0.05); 0.012 | 0.077 (0.031); 0.012 | -0.12 (0.30); 0.69 | -0.048 (0.20); 0.81 |
| UKB + MVP meta-analysis | 0.088 (0.04); 0.039 | 0.075 (0.026); 0.0042 | 0.012 (0.19); 0.95 | 0.040 (0.12); 0.74 |

Detailed results for all primary analyses can be found in the supplementary material (supplementary figures 1-16).

### Cross-population meta-analyses

We conducted a cross-population meta-analysis incorporating both the EUR and AFR GWAS described previously, as well as data from smaller GWAS of EAS and AMR participants from both cohorts (see supplementary material for results of these primary GWAS). The results of both short versus normal and long versus normal sleep duration were filtered to remove any variant that was not present across all four population groups in at least one of the primary cohorts.

After filtering, the analysis of short (n=62,102) versus normal (n=414,588) sleep duration meta-analysis resulted in a total of 7,574,717 imputed genetic variants, among which we identified 84 independent GWS risk loci (Figure 2, supplementary tables 7 and 8).

**Figure 2.**
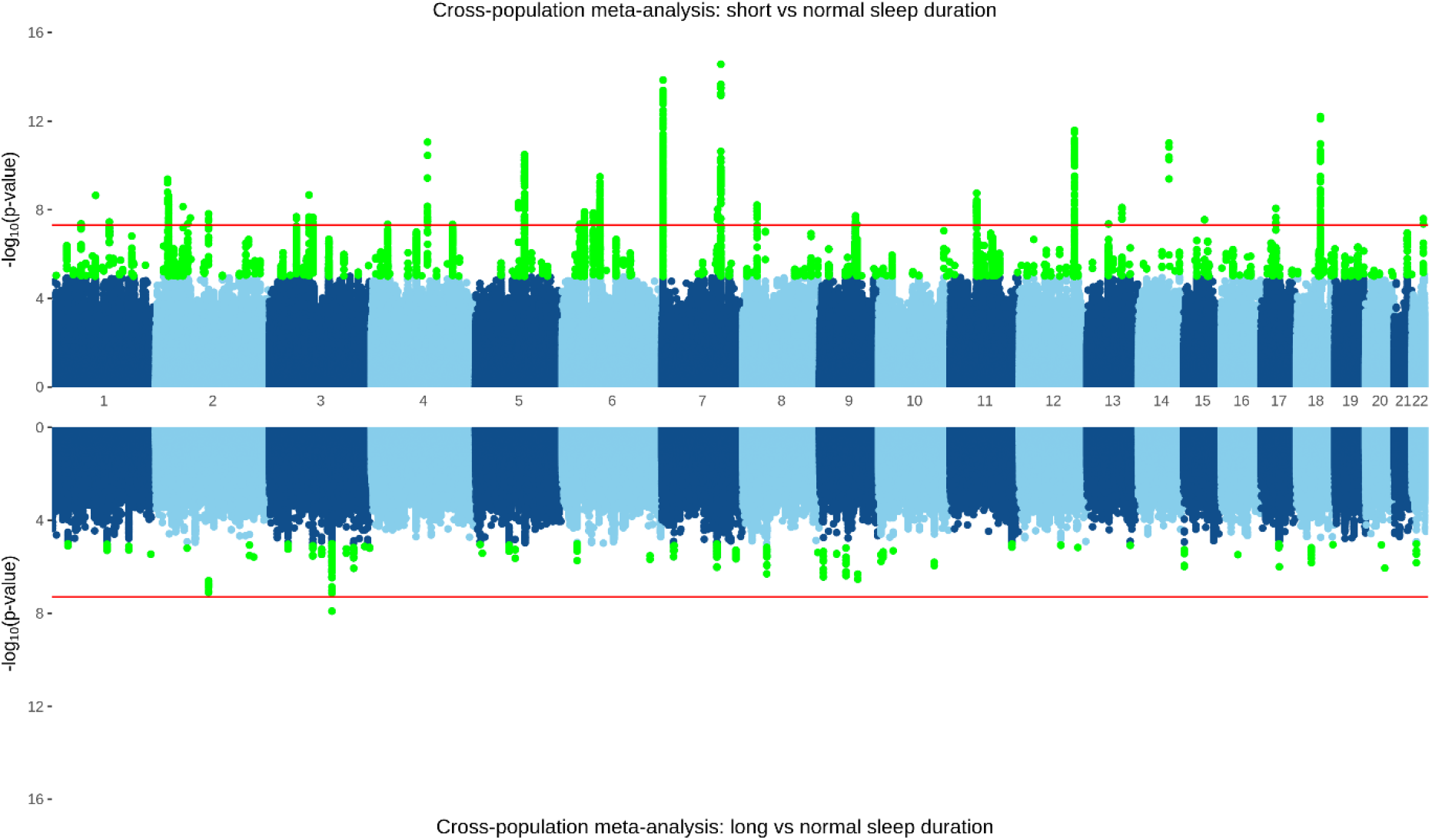
A cross-population meta-analysis including EUR and AFR participants from both UK Biobank and MVP. TOP: short (≤5 hours, n=47,928) versus normal (7-8 hours, n=453,026) sleep duration, with 28 independent genetic-risk loci reaching genome-wide significance highlighted in green. BOTTOM: long (≥10 hours, n=11,525) versus normal sleep duration with one genome-wide significant locus.

After filtering, the meta-analysis of long (n=17,715) versus normal sleep duration resulted in a total of 7,282,278 imputed genetic variants and revealed one genome-wide significant association on chromosome 3 (rs9810253, OR=1.11±0.02, p=1.24×10^−8^) (Figure 2, supplementary tables 7 and 8). Several of the top loci for both long and short sleep have been previously associated with sleep-related traits, as well as other neuropsychiatric traits (supplementary tables 8 and 9).

### Gene-based tests

A gene-based test mapped input SNPs for the short vs normal meta-analysis in EUR subjects to 18,565 protein-coding genes. Of these, 54 reached a Bonferroni-adjusted significance threshold of 2.69×10^−6^ (Figure 3**Error! Reference source not found**.). The top gene identified was *FOXP2* (p=2.29×10^−13^). A gene-based test mapped input SNPs for the long vs normal meta-analysis in EUR subjects to 18,460 protein-coding genes; none reached the Bonferroni-adjusted significance threshold of 2.65×10^−6^ (supplementary figure 19).

**Figure 3.**
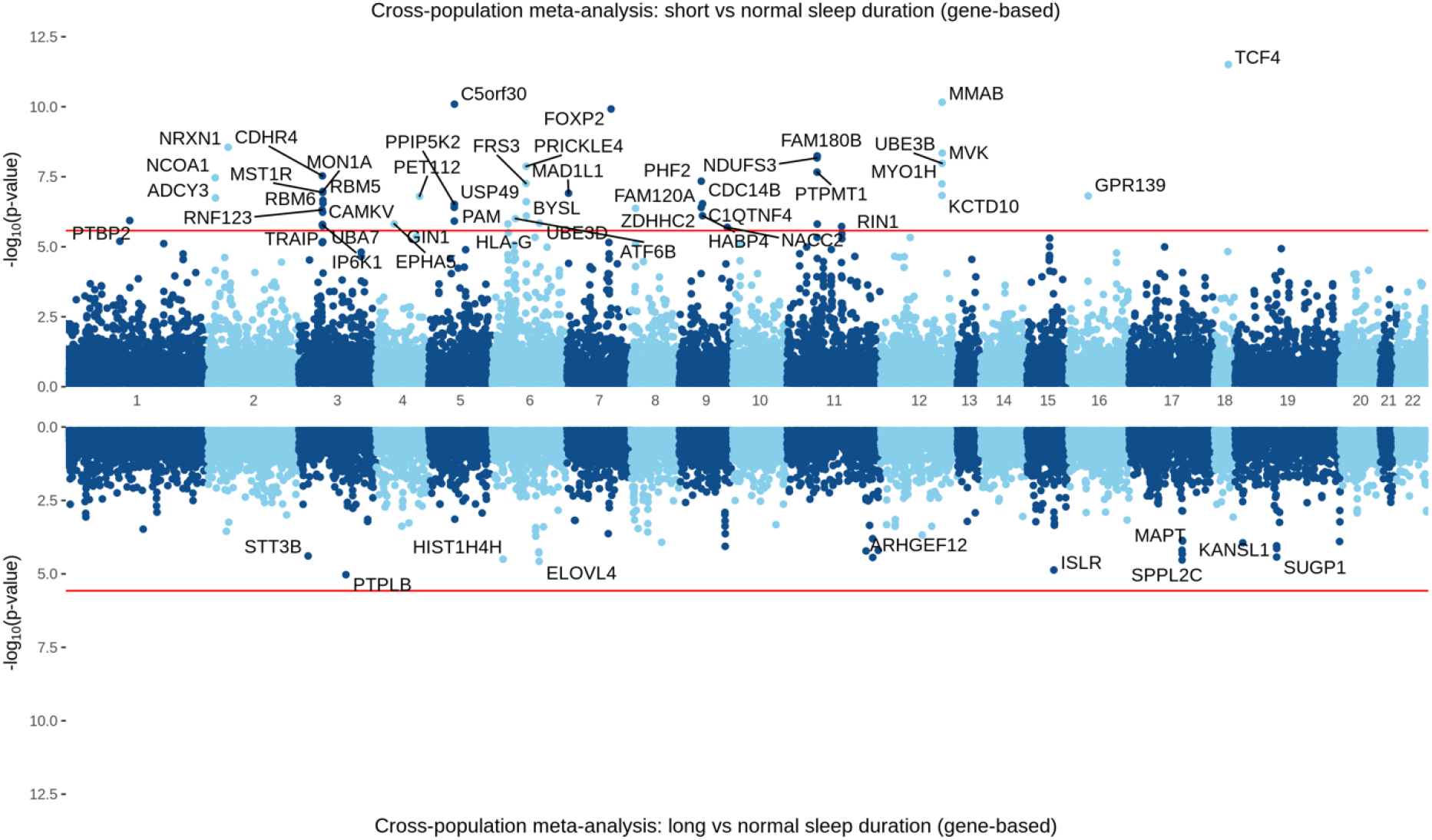
Gene-based test for short (≤5 hours, n=47,928) versus normal (7-8 hours, n=453,026) sleep duration (top) and long (≥10 hours, n=11,525) versus normal sleep duration (bottom) sleep duration in cross-population meta-analysis of UK Biobank and MVP cohorts. Bonferroni-adjusted significance threshold P<2.7×10^−6^.

**Figure 4.**
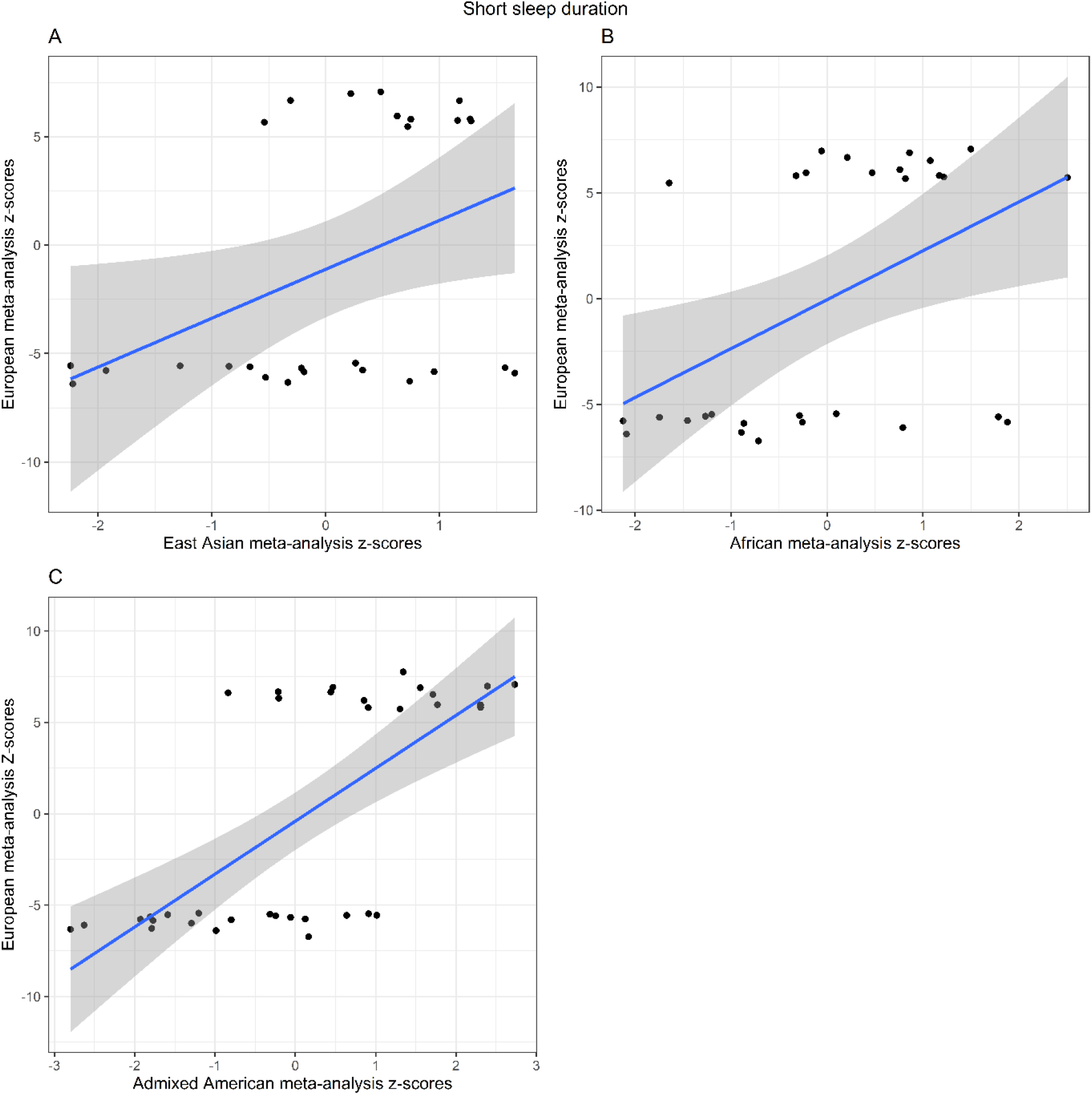
Cross-population replication analyses: Scatter plot for the z-score effect sizes for 46 genome-wide significant loci from the EUR meta-analysis on short sleep on the y axis against the z-score effect sizes for the same loci in a) EAS-only meta-analysis (ρ=0.290), b) AFR-only meta-analysis (ρ=0.293), c) AMR-only meta-analysis (ρ=0.398).

In the AFR gene-based test, input SNPs for the short vs normal meta-analysis mapped to 18,292 protein-coding genes, none of which reached a Bonferroni-adjusted significance threshold of 2.73×10^−^ 6 (supplementary figure 21). Input SNPs for the long vs normal meta-analysis mapped to 19,074 protein-coding genes; none reached a Bonferroni-adjusted significance threshold of 2.62×10^−6^.

For the cross-population meta-analysis of short sleep duration, in a gene-based test, input SNPs for the short vs normal analysis mapped to 18,914 protein-coding genes. Of these, 47 reached a Bonferroni-adjusted significance threshold of 2.66×10^−6^. The top gene identified was *TCF4* (p=53.11×10^−12^) (Figure 3). For long sleep duration, input SNPs for the long vs normal analysis mapped to 18,903 protein-coding genes; none reached a Bonferroni-adjusted significance threshold of 2.67×10^−6^ (Figure 3).

### Cross-population transferability of loci

We performed cross-population lookups for the identified ancestry-specific GWS loci across the summary statistics for EUR, AFR, and EAS populations. Of the 46 SNPs that reached genome-wide significance in the EUR meta-analysis of short sleep duration, 29 were present in the AFR summary statistics and 19 of them had a direction of effect consistent with the EUR results. Three of these loci reached nominal significance of p < 0.05 (rs12705972 on chromosome 7, p= 0.012; rs2111216 on chromosome 12, p= 0.03; rs7313797 on chromosome 12, p= 0.04)

In the EAS summary statistics, 27 of the EUR GWS loci were present and 19 had a consistent direction of effect. Two of these loci reached nominal significance of p < 0.05 (rs146618518 on chromosome five, p= 0.025; rs7313797 on chromosome 12, p= 0.026).

Twenty-two of the 46 EUR GWS loci were present in both AFR and EAS, and 10 of these had a consistent direction of effect across populations. Of these 10, only one SNP reached nominal significance in all three studies (rs7313797 on chromosome 12) (supplementary table 11).

Rs62158206, the only SNP reaching genome-wide significance for long sleep duration in the EUR meta-analysis, was present in both the AFR (same direction of effect) and EAS summary statistics (opposite direction of effect), but P > 0.05 in both cases.

Given the known differences in LD structure and allele frequency across the population groups, we also considered all SNPs in high LD (r^2^ > 0.6) with the GWS loci from the EUR meta-analysis of short sleep. One SNP was successfully matched to three SNPs in high LD that reached nominal significance in the AFR ancestry results (query SNP rs62144584, matched to a total of 14 SNPs in the AFR population with P<0.05). The majority (11 out of 14) of these matched SNPs show a consistent direction of effect with the EUR results. Another SNP, rs201640077, was matched to three SNPs in the EAS population that reached nominal significance, with a consistent direction of effect (supplementary table 12).

### Genetic correlation analysis

The within-phenotype genetic correlation between EUR participants in UK Biobank and MVP cohorts is 0.84 (±0.05, p=3.74×10^−87^) for short sleep and 1.106 (±0.17, P=1.31×10^−10^) for long sleep. For AFR, the correlations were non-significant for short sleep and long sleep could not be calculated for long sleep due to low sample sizes (supplementary table 13). There was no significant cross-ancestry genetic correlation between the EUR and AFR participants for either short or long sleep (supplementary table 13).

We estimated a r_g_ between short and long sleep of 0.16 (±0.04), p=2.0×10^−4^ in the EUR-only meta-analyses and -0.09 (±0.1), p=0.72 in the AFR-only meta-analyses. The weak genetic correlation in the EUR sample suggests a distinct genetic architecture underlying the two traits. The lack of significant genetic correlation between these traits in the AFR sample may be due to lack of power, given the lower sample size in this analysis (supplementary table 15).

LDSC analysis was conducted in the EUR sample only (Figure 5), due to lack of available comparator data for AFR participants. The traits most strongly associated with both short and long sleep duration were years of schooling (short: r_g_ = -0.48 (0.02), p=2.41×10^−105^; long: r_g_ = -0.36 (0.03), p=2.18×10^−24^) and sleep duration (as a continuous trait), with directions of effect consistent with the definitions of short and long sleep duration (short: r_g_ = -0.76 (0.04) p=6.05×10^−72^; long: r_g_ = 0.39 (0.06), p=1.99×10^−11^). In addition, short sleep duration was positively correlated with attention deficit hyperactivity disorder (r_g_ = 0.53 (0.14), p=1.0×10^−4^) and negatively correlated to bipolar disorder (r_g_ = -0.16 (0.04), p=2.0×10^−4^). Long sleep duration was also positively correlated to schizophrenia (r_g_ = 0.30 (0.05), p=1.58×10^−10^).

**Figure 5.**
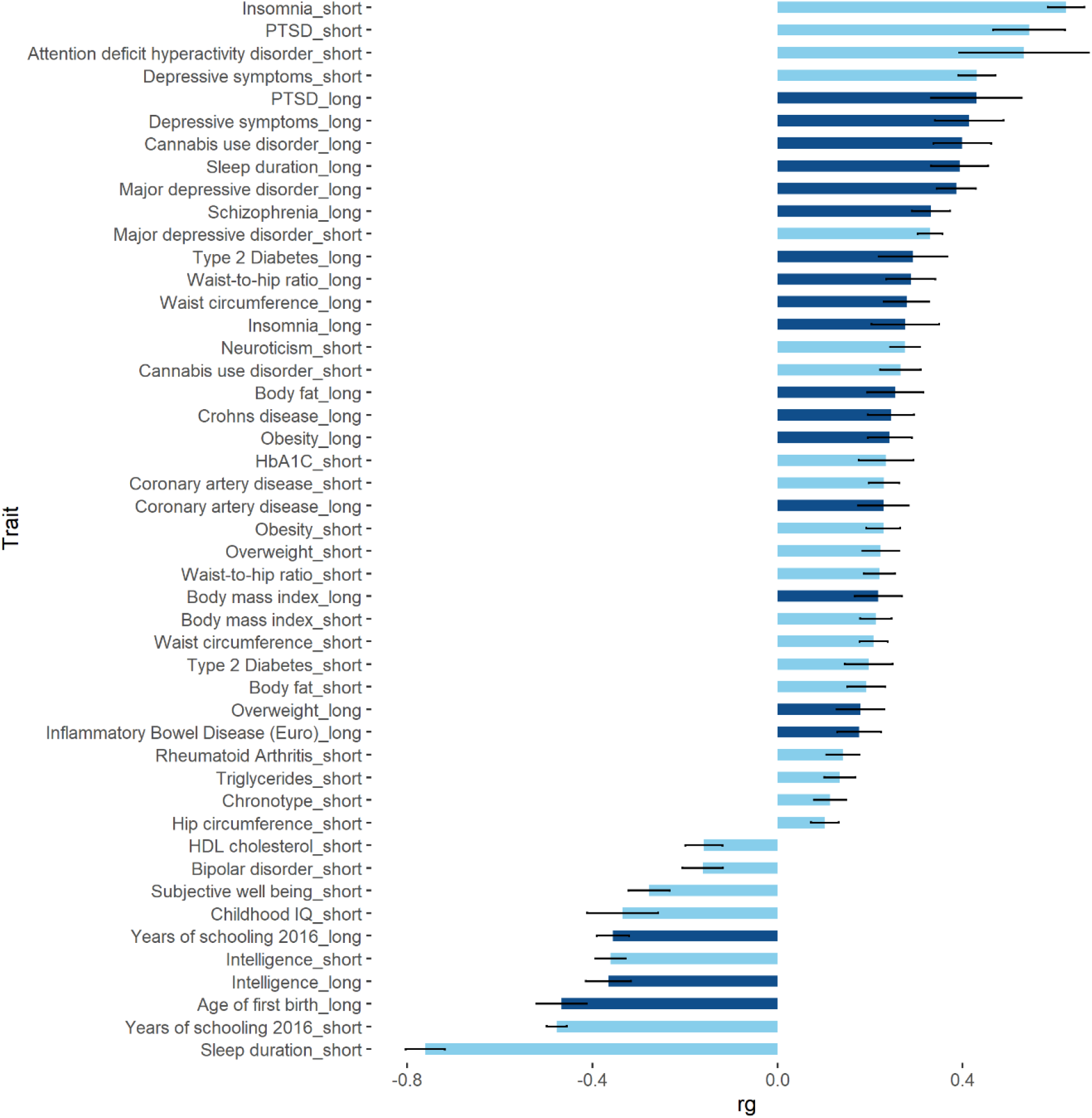
Summary of significant (Bonferroni correction for 40 tests; P<1.25×10^−3^) genetic correlations between short (light blue) and long (dark blue) sleep duration and previously published GWAS results (EUR only).

We observed several significant correlations between sleep duration and cardiometabolic traits. For example, both short and long sleep were positively correlated with coronary artery disease (short: r_g_ = 0.23 (0.03), p=1.8×10^−12^; long: r_g_ = 0.23 (0.05), p=2.6×10^−5^), obesity (short: r_g_ = 0.23 (0.04), p=3.2×10^−10^; long: r_g_ = 0.24 (0.05), p=4.2×10^−7^), and type 2 diabetes (short: r_g_ = 0.20 (0.05), p=1.0×10^−4^; long: r_g_ = 0.29 (0.07), p=7.8×10^−5^). Additional significant (Bonferroni correction for 40 independent traits; p<1.25×10^−3^) correlations are summarised in Figure 5 and in the supplementary table 15.

### Mendelian Randomisation

MR analysis investigating the causal influence of short sleep on depression supported a positive causal association between short sleep and increased risk of depression, using the inverse variance weighted method (β=0.19 (0.02) p=1.5×10^−19^, Figure 6, supplementary table 16). Conversely, MR on the impact of depression on short sleep did not support causal association (β=0.01 (0.03) p=0.69). MR investigating the causal influence of long sleep on depression reveals a positive causal association between long sleep and increased risk of depression, using the inverse variance weighted method (β=0.14 (0.03) p=1.64×10^−5^). This was a bidirectional effect, with an analysis of the impact of depression on long sleep also revealing evidence of a causal association (β=0.14 (0.04) p=7.6×10^−5^).

**Figure 6.**
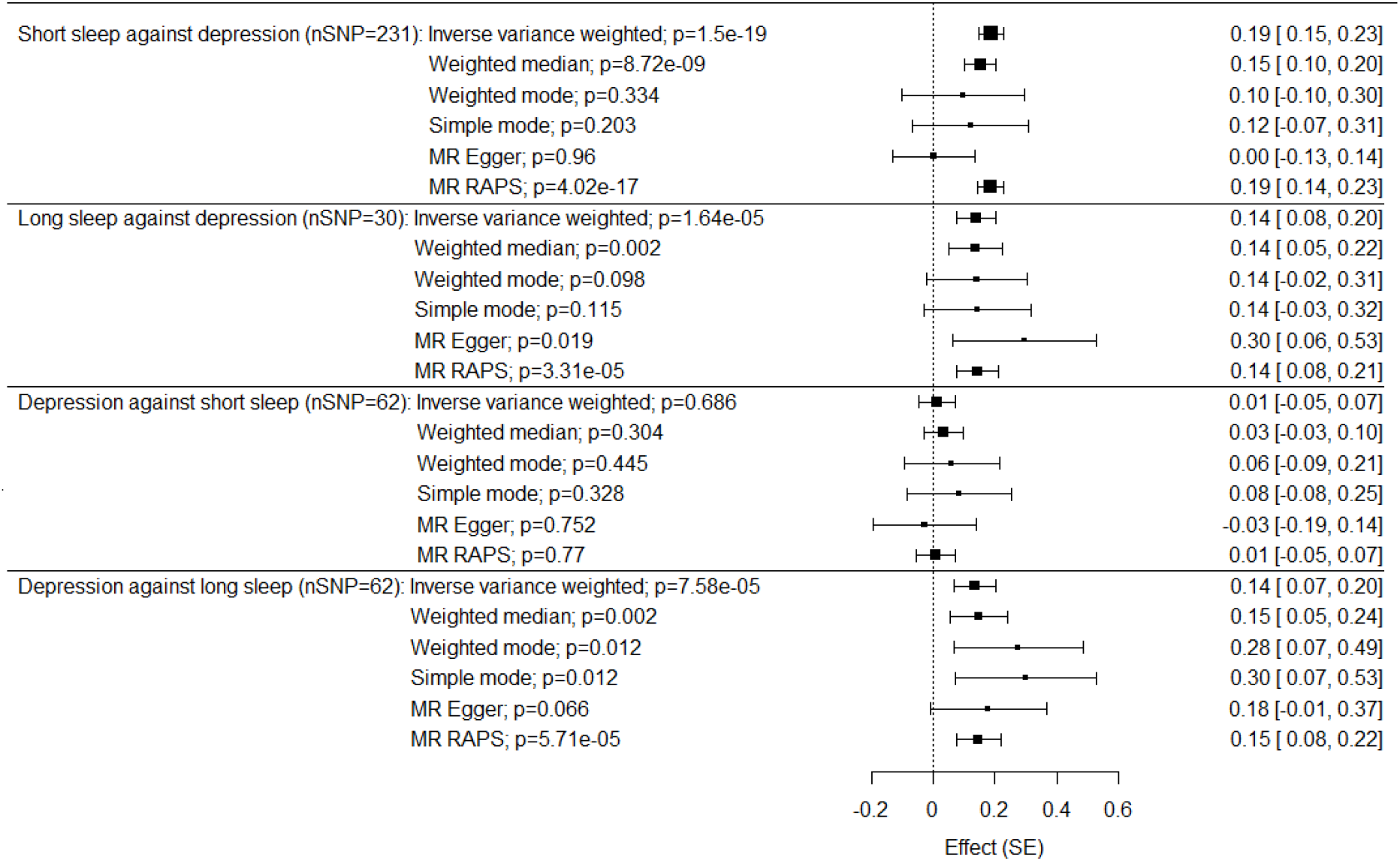
Effect estimates for the bi-directional Mendelian randomisation analyses between short and long sleep and depression. The forest plots show the estimates and 95% confidence intervals for the association between exposures and outcomes using different MR methods.

The Egger-intercept was non-significant in three of four analyses (long sleep against depression: Egger intercept -0.01 (0.009), p=0.18; depression against short sleep: Egger intercept 0.003 (0.005), p=0.62; depression against long sleep: Egger intercept -0.003 (0.006), p=0.63). The Egger intercept for short sleep against depression was significant, (0.009 (0.003), p=0.004), suggesting the inverse variance weighted estimate may be biased. We therefore repeated this analysis using a more stringent p-value threshold for the exposure SNPs of 5×10^−8^. In this case, the Egger intercept was non-significant (0.005 (0.012), p=0.65) the inverse variance weighted estimate remained significant (0.23 (0.046), p=4.0×10^−7^).

Weighted median, MR-Egger, and MR-RAPS analyses were conducted as sensitivity analyses. The weighted median regression and MR-RAPS analyses were significant for depression against long sleep, and both short and long sleep against depression, following the same pattern as observed using the inverse-variance weighted method. The MR-Egger analyses revealed only one significant association, for long sleep against depression.

## Discussion

The question of how to improve sleep quality and optimize sleep duration is of constant interest, with the global market for sleep aids and technologies exceeding 80 billion US-dollars per year (51). Along with genetic influences, a wide variety of demographic, social, and environmental factors can impact quality and duration of sleep, including socioeconomic status, stressful life events, home and neighbourhood characteristics, work and school schedules, medication and substance use, and mental and physical health conditions. Indeed, sleep quality and duration can be considered both a risk factor for and symptom of many health conditions.

We present a large GWAS of self-reported sleep duration conducted for the first time in a diverse population. In addition to replicating associations with many genes previously linked to sleep traits, both in studies with overlapping samples and fully independent cohorts, our analyses expand on the previous understanding of the genetic architecture of sleep through the identification of numerous novel risk loci for short sleep duration and one novel locus for long sleep duration. We identify genes of interest in EUR, AFR, and cross-population analyses. Our findings add to the existing knowledge of the genetic basis of sleep duration, as well as highlighting ancestry-specific risk loci and shared genetic risk with a variety of cognitive, neuropsychiatric, and metabolic traits.

We conducted GWAS separately for individuals of EUR and AFR ancestry before conducting a cross-population meta-analysis that also incorporated EAS and AMR samples. In a GWAS of short vs normal sleep duration in the UK Biobank and MVP EUR samples, we identify 46 independent GWS risk loci. The strongest associations can be found on chromosome 7 (rs6466488, near *FOXP2* and rs111595851, near *MAD1L1)* and chromosome 4 (rs13107325, near *SLC39A8*). *FOXP2* is a transcription factor which has been implicated in GWAS of insomnia, BMI, cannabis use disorder, and risk-taking, as well as short sleep duration (7,10,15,52). *MAD1L1* is a member of a family of genes that encode proteins important in the mitotic checkpoint. This gene has previously been associated with sleep and several psychiatric traits in previous GWAS, including bipolar disorder, anxiety, PTSD, major depressive disorder, and schizophrenia (7,53–55). *SLC39A8* (also known as *ZIP8*) is a transmembrane protein, with a role in zinc transportation. Genetic polymorphisms in *SLC39A8* have been associated with alcohol consumption, schizophrenia, and insomnia (15,56,57). A MAGMA gene-based test reveals 37 significant genes (p<2.6×10^−6^), with the top association at *TCF4* (p=2.29×10^−13^), a transcription factor that has previously been associated with cognitive traits, educational attainment, alcohol consumption, autism spectrum disorder, schizophrenia, depression, lung function and BMI, as well as sleep duration (15,46,57–61).

In EUR, we identified one locus with compelling evidence that it results in long sleep duration. The strongest association was at rs62158206 on chromosome 2. This SNP is intergenic to *PAX8*, which encodes a transcription factor that has previously been associated with several sleep related phenotypes, including insomnia (with a consistent and opposite direction of effect) (15,16) and sleep disturbance in depression (62). In addition, this gene has been highlighted in several previous GWAS on sleep duration, both in fully independent and overlapping samples (7,8). SNP-based heritability of short sleep duration was 11.9% (p=2.45×10-115). Long sleep duration appeared less heritable, with SNP-based heritability of 7.8% (p=1.61×10-20). Both estimates are broadly consistent with previously published results (7,8). The significant difference in the number of significant loci for long sleep compared to short sleep may be in part a result of this lower heritability, perhaps suggesting a greater influence of environmental factors on longer sleep duration; but lower power based on sample size may have been decisive for both measures (risk loci and observed heritability). There were also fewer long sleepers (≥10 hours) in both the UK Biobank and MVP cohorts compared to short sleepers (≤6 hours) and the effect estimates for the top associations were of comparable magnitude, indicating decreased power for the analysis.

Amongst AFR participants, we identified one risk locus, at rs148926968 on chromosome 13 clearly associated with long sleep. This locus is intergenic for *LMO7*, which has been previously identified in GWAS of obsessive-compulsive disorder (63), several age-related diseases including hearing loss (64) and Alzheimer’s disease (65), and several cross-population GWAS on eyesight-related traits (66,67). In AFR, we found significant SNP-based heritability for short sleep duration (8.2%, p=0.04), but not for long sleep duration. There are no prior published h^2^ estimates for sleep duration in individuals of AFR. Given the lower observed heritability of long sleep in EUR, we suspect that the sample size included here is too small to confirm heritability in AFR.

We conducted a cross-population meta-analysis including all the EUR and AFR data previously described, as well as additional EAS and AMR cohorts from UK Biobank and MVP. We identified 84 independent GWS risk loci for short sleep duration. The strongest associations were on chromosomes 7 (rs1989903, near *FOXP2*, and 7:2054314:C:CG, near *MAD1L1*) and 18 (rs11152363, near *TCF4*).

The gene-based test identified 47 significant genes, with the strongest association with *TCF4*. Several of these significant loci and genes are the same as highlighted in EUR-only meta-analysis, which is unsurprising given the relative sample sizes of each population group. However, the increased number of genome-wide significant associations shows how the diverse sample increased power and added valuable information.

In the cross-population analysis of long sleep duration, we identified one GWS locus at rs9810253 on chromosome 3 (p=1.24×10^−8^). This locus has a consistent direction of effect across all primary studies, with p=7.36×10^−7^ in the EUR-only analysis and 0.005 in the AMR analysis.

Due to the limited GWS findings in the non-EUR analyses, we performed cross-population lookups in the EUR meta-analysis of UK Biobank and MVP cohorts. Of the 46 GWS loci in this EUR study, 22 were also present in AFR and EAS meta-analysis, and ten had a consistent direction of effect across all three populations. Of these ten, only one reached at least nominal significance in all populations: rs7313797 on chromosome 12. This SNP is intronic to *KCTD10*, which has previously been identified in GWAS of neurotic disorder, well-being, coronary artery disease, and HDL cholesterol levels (68– 71).

Our results do not support significant genetic correlation between the EUR and AFR samples for either short or long sleep, though these analyses are hindered by small sample sizes in the AFR population. We do find some evidence for portability across populations, as highlighted in Figure 4, but future analysis in additional non-EUR datasets and in a larger AFR sample should help clarify the extent of genetic overlap and aid the identification of truly causal variants.

The genetic correlation between short and long sleep was low, with r_g_=0.16 (p=0.0002) in EUR and a non-significant correlation in AFR, due to the smaller sample size. Larger samples are necessary to further evaluate the relationship of these sleep traits in non-European populations. Nevertheless, the result in the EUR analysis confirms our hypothesis that these traits, though clearly phenotypically related, possess distinct underlying genetic architecture and biology. There are notable similarities for some traits significantly correlated with both short and long sleep, such as depression, cannabis use disorder, PTSD, coronary artery disease, obesity, and diabetes. This indicates that an increased genetic risk for a range of traits is associated with increased risk of sleep disturbance at either end of the spectrum, perhaps depending on the specific variants at play. In addition, short and long sleep were both significantly positively correlated with insomnia. Localised genetic correlation analyses may be valuable in establishing if these shared patterns of r_g_ are in fact driven by the same loci (72).

There is significant evidence, genetic and otherwise, of high comorbidity between disturbed sleeping and a variety of neuropsychiatric, cognitive, and metabolic traits. Short and long sleep duration have been associated with all-cause mortality and decreased life expectancy. The results of the genetic correlation analyses in LDSC provide support for these observations and confirm that some of this comorbidity can be explained by shared genetic risk.

The MR analyses found a directional casual influence of short sleep duration resulting in increased risk of depression. We also identified evidence of a bi-directional causal relationship between long sleep duration and depression. Many previous studies have identified phenotypic association between sleep duration and depression (73–77), but the causal direction of these associations has not always been clear. The findings from the present analyses, that both short and long sleep duration can cause an increased risk of depression, supports existing theories on the importance of healthy sleep patterns for mood regulation and highlights shared genetic risk factors between the two extremes.

We considered hours of daylight exposure as a potential environmental factor impacting variance in sleep duration. In the UK Biobank sample, where we were able to consider monthly solar irradiation data, higher levels of estimated solar irradiation were significantly associated with shorter reported sleep duration. In the MVP sample, where we assessed annual irradiation only, increased solar irradiation was not significantly associated with sleep duration. Independently of solar irradiation levels, we found that month of recruitment was significantly associated with sleep duration.

It is possible that the use of annual data only in the MVP sample may be limiting our ability to detect an effect; the large variation in daylight exposure across the year cannot be captured by annual data. However, there are other significant differences between the cohorts that could contribute to these findings, and the question on sleep duration in both UK Biobank and MVP was phrased such as to ask for habitual sleep patterns. Our data suggests solar irradiation is a significant influence, however longitudinal data recording sleep patterns over the course of several years will be valuable in determining the full extent of the impact of hours of daylight on sleep duration.

Reliance on self-reported sleep data is a limitation. However, a previous study in the UK Biobank demonstrated a high level of consistency between the top variants identified in a GWAS of self-report sleep duration data and those seen in a study of 85,499 subjects using wrist-worn accelerometer data (7). Both cohorts included predominantly older adults, where sleep disturbance (particularly short sleep) is more common and while this increases the power for the analyses intended, it limits our ability to generalise findings to younger groups. A further limitation is in the underrepresentation of non-European populations. Although we were able to include four population groups in the cross-population meta-analysis, many post-GWAS analyses were conducted in EUR and AFR ancestry groups only, and some of the methods used are less reliable in population groups with higher levels of admixture.

In summary, we have identified multiple novel variants of GWS for short sleep duration, and one novel locus for long sleep duration, among UK Biobank and MVP cohorts. Several of these loci and genes that warrant future investigation in populations of greater age, sex, and ancestral diversity. Genetic correlations provided support for shared genetic risk between sleep duration and a range of comorbid traits and MR analysis supports a causal association between sleep duration and depression. These findings highlight the value of understanding the genetic basis of sleep patterns in order to improve public health.

## Data Availability

Summary statistics will be made available on dbGap by the Million Veteran Program following publications.

https://www.ncbi.nlm.nih.gov/projects/gap/cgi-bin/study.cgi?study_id=phs001672.v8.p1

## Acknowledgements

This research has been conducted using the UK Biobank (www.ukbiobank.ac.uk) under application number 8901 and 11362 (PI: Andrew McQuillin, Co-I: Elvira Bramon). This research is also based on data from the Million Veteran Program, Office of Research and Development, Veterans Health Administration. This publication does not represent the views of the Department of Veteran Affairs or the United States Government. This work is supported by funding from the UK Medical Research Council and the US Department of Veteran Affairs (CSP575b (NCT02256644) and MERIT (I01CX001849) grants). Daniel F. Levey was supported by a NARSAD Young Investigator Award from the Brain & Behavior Research Foundation and a Career Development Award from the Veterans Health Administration Office of Research and Development (Grant IK2BX005058) and is Aimee Mann Fellow of Psychiatric Genetics at Yale. Joseph D. Deak was supported by the National Institute on Alcohol Abuse and Alcoholism (NIAAA) T32 AA028259. Haritz Irizar has received funding from the European Union’s Horizon 2020 research and innovation programme under the Marie Sklodowska-Curie grant agreement no 747429 and is currently supported by a grant from the National Institute of Allergy and Infectious Diseases, National Institutes of Health. Elvira Bramon acknowledges the Medical Research Council UK (MR/W020238/1, G0901310, G1100583 and G1100583), the Wellcome Trust (085475/B/08/Z, 085475/Z/08/Z), National Institute of Health Research UK (NIHR200756) and the NIHR Biomedical Research Centre at University College London Hospitals.

The authors thank all the volunteers who participated in UK Biobank and the Million Veteran Programme.

## LDHub

We gratefully acknowledge all the studies and databases that made GWAS summary data available: **ADIPOGen** (Adiponectin genetics consortium), **C4D** (Coronary Artery Disease Genetics Consortium), **CARDIoGRAM** (Coronary ARtery DIsease Genome wide Replication and Meta-analysis), **CKDGen** (Chronic Kidney Disease Genetics consortium), **dbGAP** (database of Genotypes and Phenotypes), **DIAGRAM** (DIAbetes Genetics Replication And Meta-analysis), **ENIGMA** (Enhancing Neuro Imaging Genetics through Meta Analysis), **EAGLE** (EArly Genetics & Lifecourse Epidemiology Eczema Consortium, excluding 23andMe), **EGG** (Early Growth Genetics Consortium), **GABRIEL** (A Multidisciplinary Study to Identify the Genetic and Environmental Causes of Asthma in the EUR Community), **GCAN** (Genetic Consortium for Anorexia Nervosa), **GEFOS** (GEnetic Factors for OSteoporosis Consortium), **GIANT** (Genetic Investigation of ANthropometric Traits), **GIS** (Genetics of Iron Status consortium), **GLGC** (Global Lipids Genetics Consortium), **GPC** (Genetics of Personality Consortium), **GUGC** (Global Urate and Gout consortium), **HaemGen** (haemotological and platelet traits genetics consortium), **HRgene** (Heart Rate consortium), **IIBDGC** (International Inflammatory Bowel Disease Genetics Consortium), **ILCCO** (International Lung Cancer Consortium), **IMSGC** (International Multiple Sclerosis Genetic Consortium), **MAGIC** (Meta-Analyses of Glucose and Insulin-related traits Consortium), **MESA** (Multi-Ethnic Study of Atherosclerosis), **PGC** (Psychiatric Genomics Consortium), **Project MinE** consortium, **ReproGen** (Reproductive Genetics Consortium), **SSGAC (**Social Science Genetics Association Consortium) and **TAG** (Tobacco and Genetics Consortium), **TRICL** (Transdisciplinary Research in Cancer of the Lung consortium), **UK Biobank**.

We gratefully acknowledge the contributions of Alkes Price (the systemic lupus erythematosus GWAS and primary biliary cirrhosis GWAS) and Johannes Kettunen (lipids metabolites GWAS).

## Author Contributions

I.A.Z, D.F.L, and J.G. had primary responsibility for the design of the study. D.F.L. E.B., and J.G. supervised the study. I.A.Z. and D.F.L. had primary responsibility for the genetic and bioinformatics analyses, with support from O.G, J.D. M.G., H.Z., H.I., K.K., A.M, R.P., and J.G. O.G., K.K., H.I., and S.D. contributed to the initial quality control and data management of the UK Biobank data. The initial manuscript was drafted by I.A.Z, D.F.L., and J.G. Manuscript contributions and interpretation of results were provided by I.A.Z, D.F.J, D.B., R.P., M.B.S., E.B., and J.G. The remaining authors contributed to other organisational or data-processing components of the study. All authors reviewed and approved the final manuscript.

## Notes

### Competing Interest Statement

The authors have declared no competing interest.

### Author Declarations

The UK Biobank study was approved by the North-West Research Ethics Committee (ref 06/MREC08/65) in accordance with the Declaration of Helsinki. Research involving the MVP in general is approved by the VA Central Institutional Review Board. All participants in both cohorts provided written informed consent.

