## Supplementary material for "Genome-wide association study investigating short and long sleep duration: a cross-population meta-analysis"

### Results from primary studies

#### Primary studies: UK Biobank

##### European-ancestry population

Supplementary Figure 1. Mirrored Manhattan plot showing genetic associations with short sleep duration (top) and long sleep duration (bottom) in European UK Biobank participants

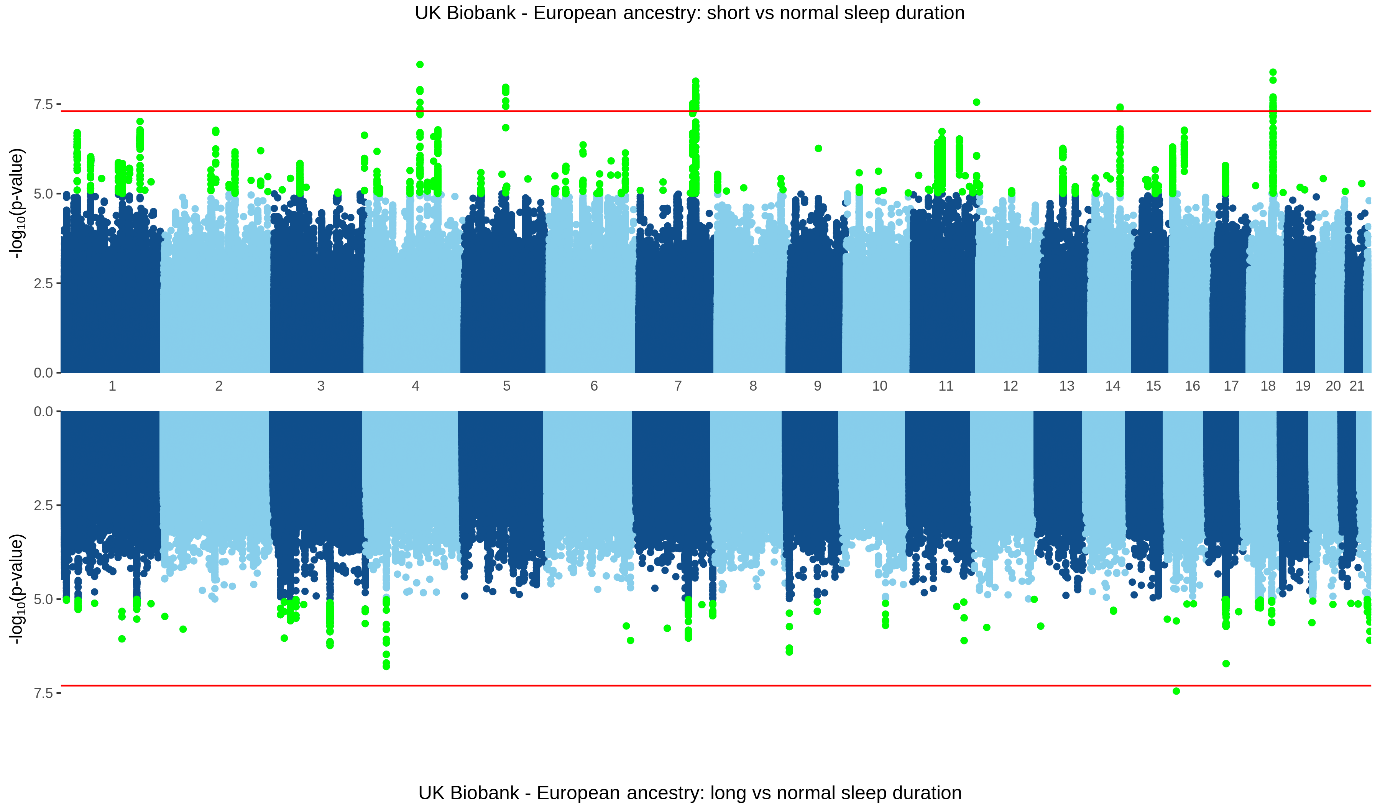

Supplementary Figure 2. Quantile-quantile plot for short sleep duration (left) and long sleep duration (right) in the European UK Biobank sample

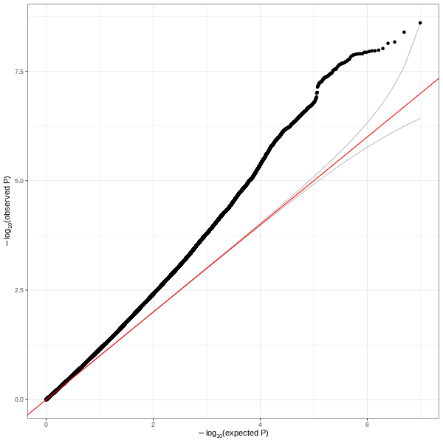

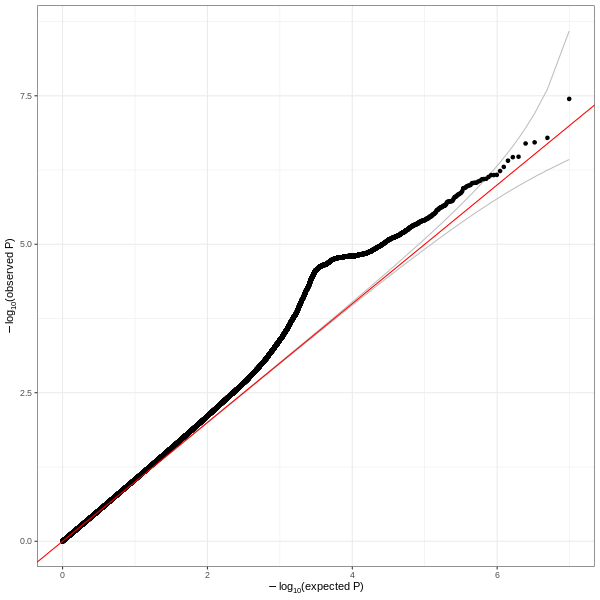

##### African-ancestry population

Supplementary Figure 3. Mirrored Manhattan plot showing genetic associations with short sleep duration (top) and long sleep duration (bottom) in African UK Biobank participants

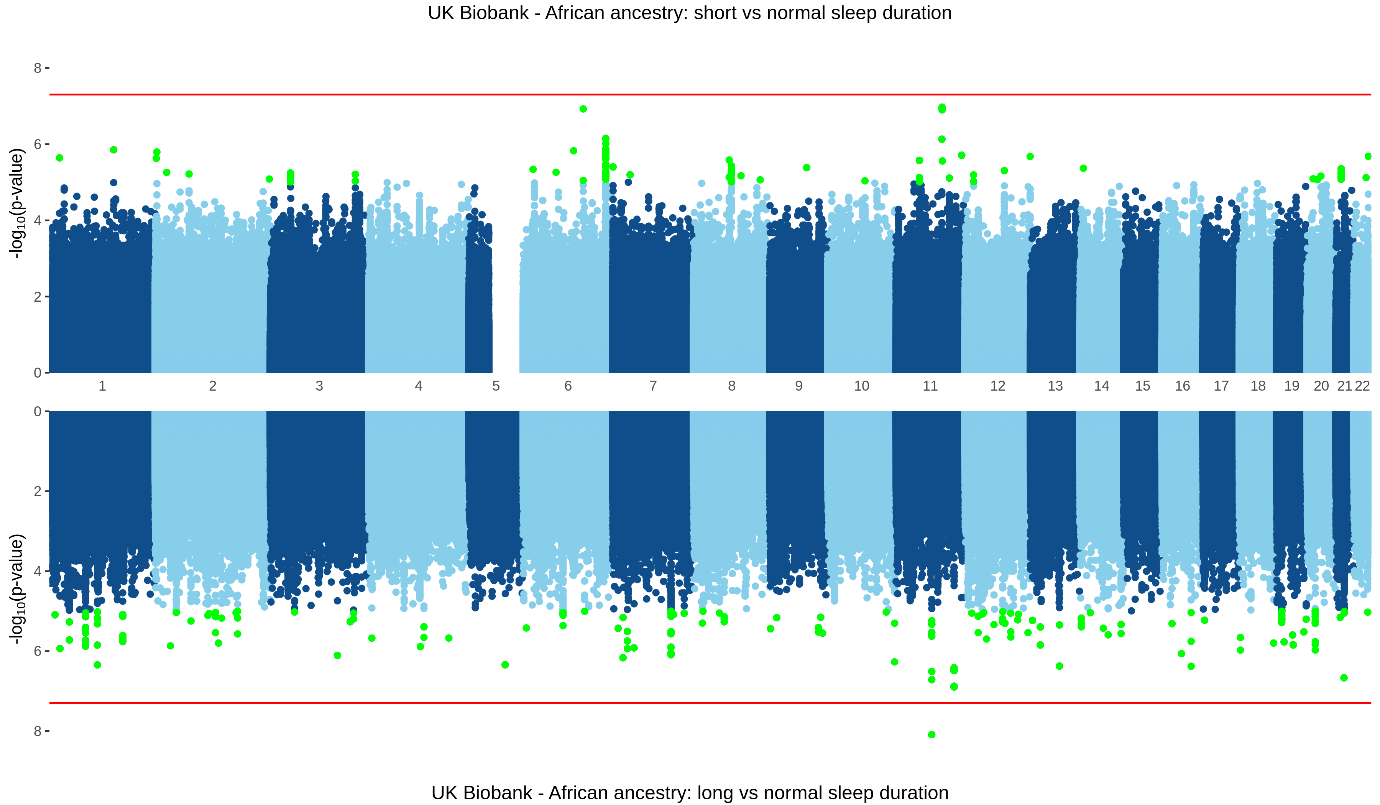

Supplementary Figure 4. Quantile-quantile plot for short sleep duration (left) and long sleep duration (right) in African UK Biobank participants

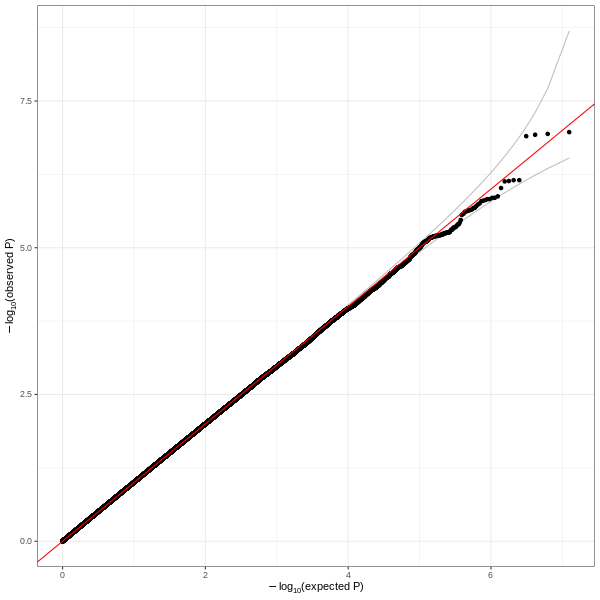

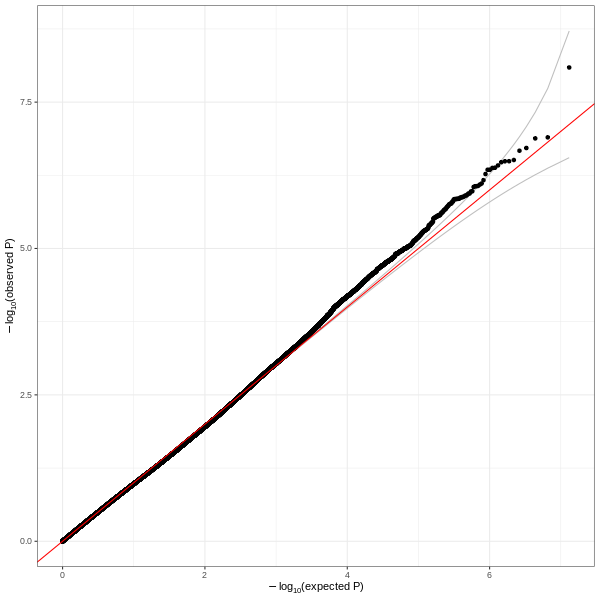

##### East Asian-ancestry population

Supplementary Figure 5. Mirrored Manhattan plot showing genetic associations with short sleep duration (top) and long sleep duration (bottom) in East Asian UK Biobank participants

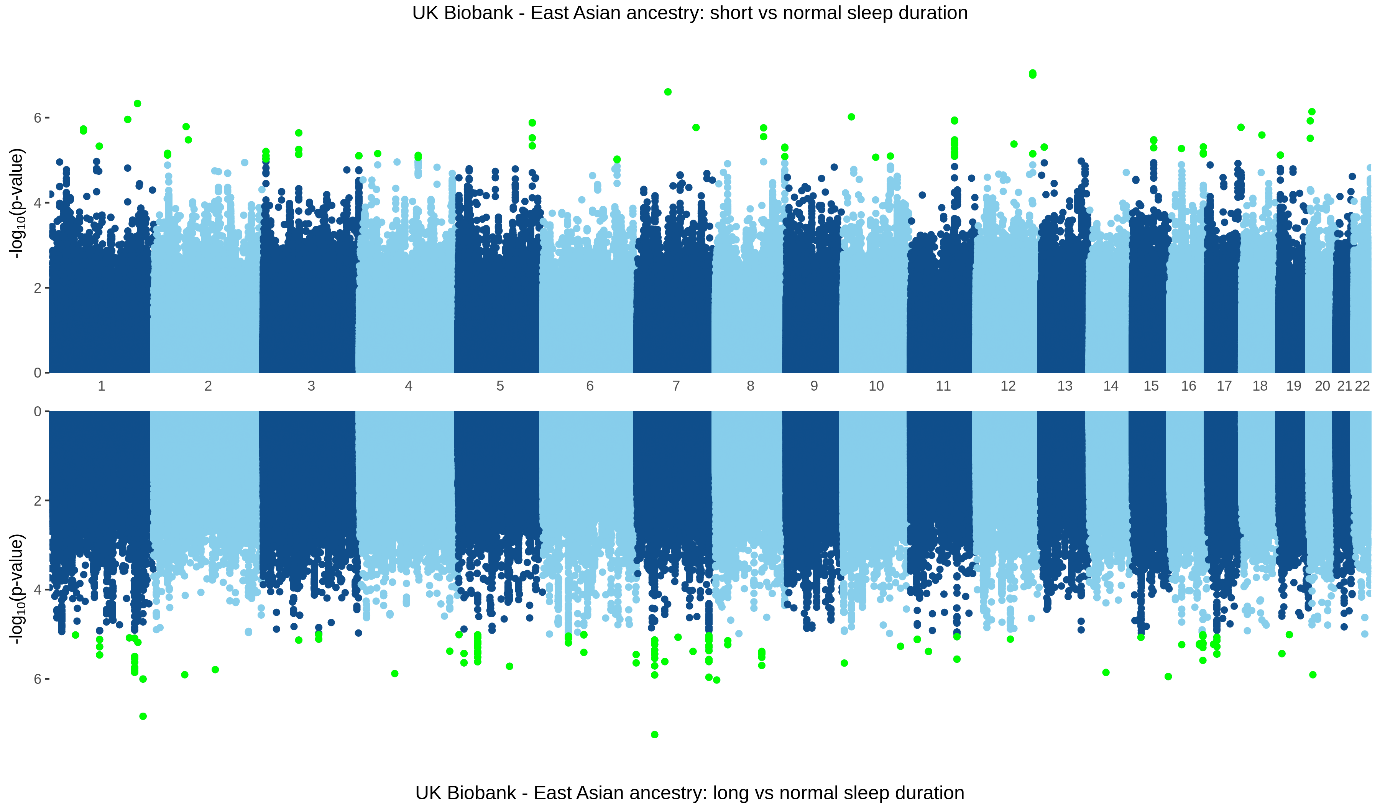

Supplementary Figure 6. Quantile-quantile plot for short sleep duration (left) and long sleep duration (right) in East Asian UK Biobank participants

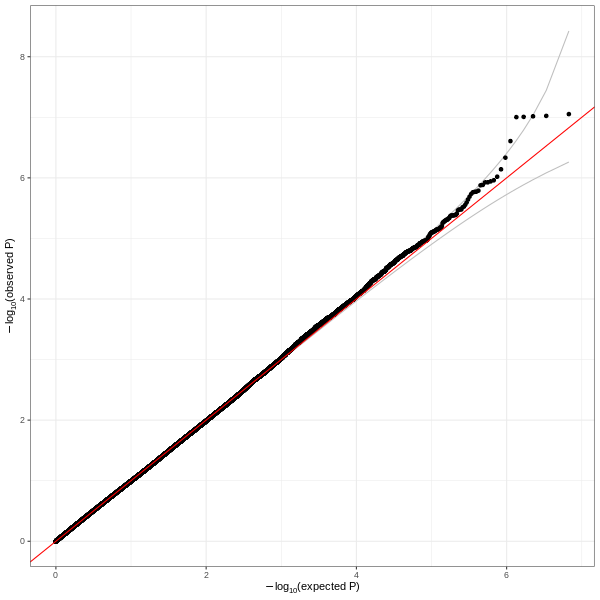

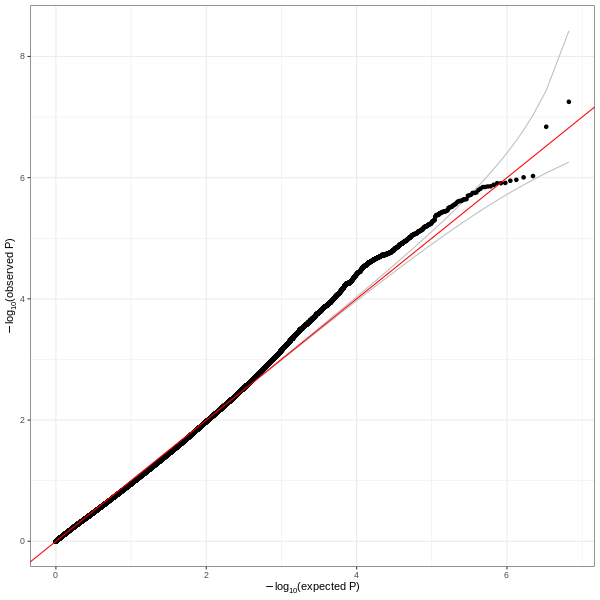

##### Admixed American population

Supplementary Figure 7. Mirrored Manhattan plot showing genetic associations with short sleep duration (top) and long sleep duration (bottom) in Admixed UK Biobank participants

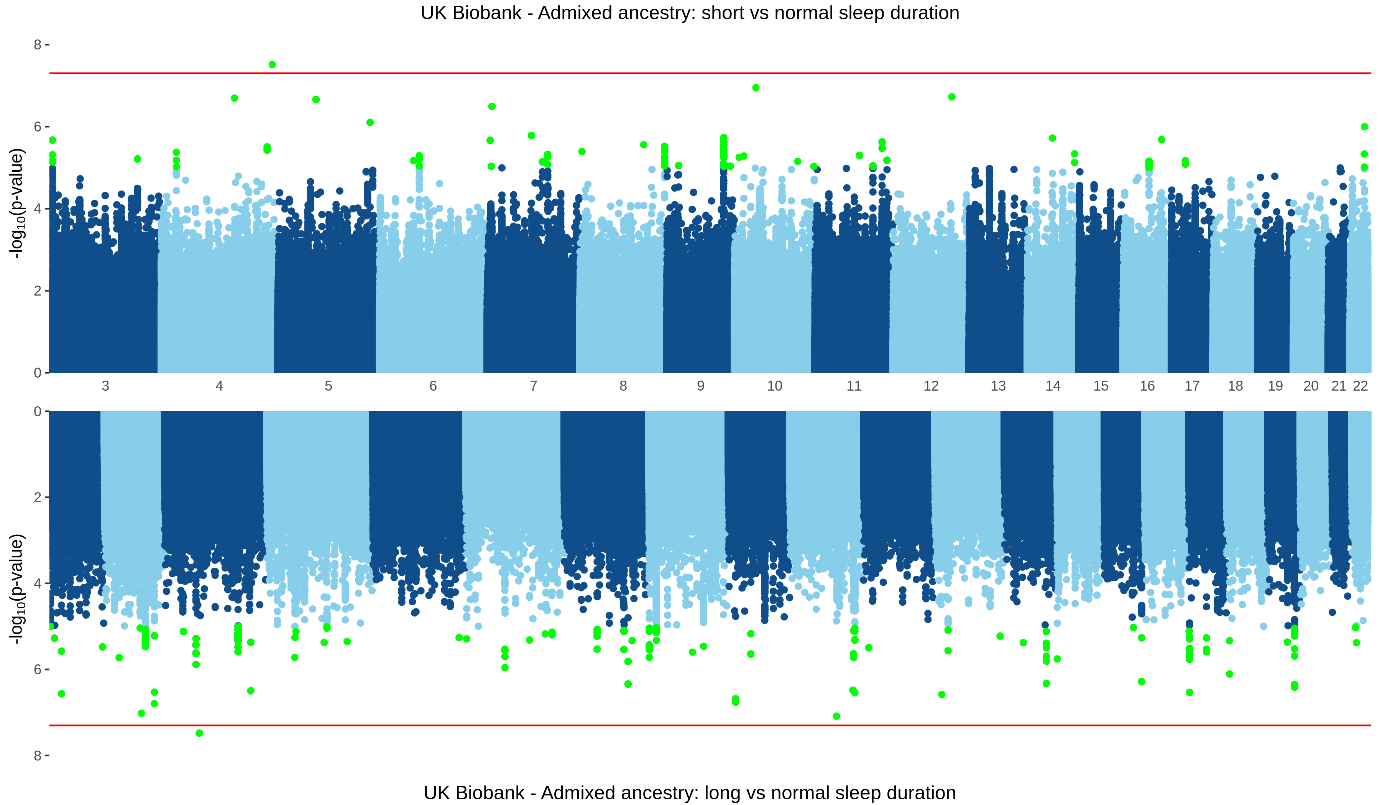

Supplementary Figure 8. Quantile-quantile plot for short sleep duration (left) and long sleep duration (right) in Admixed UK Biobank participants

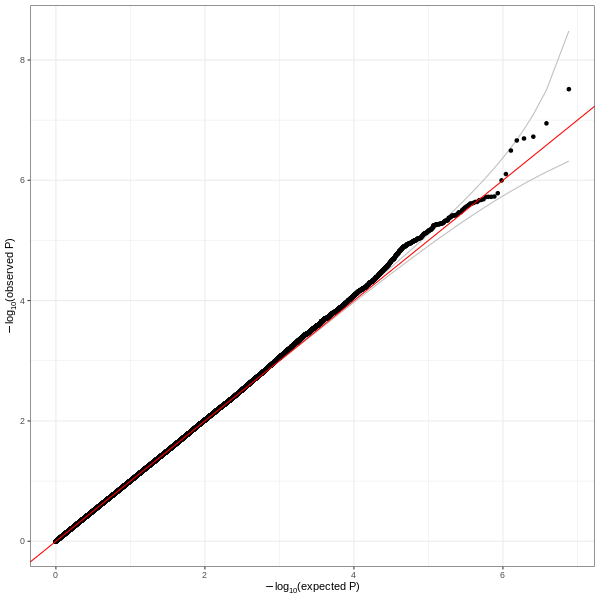

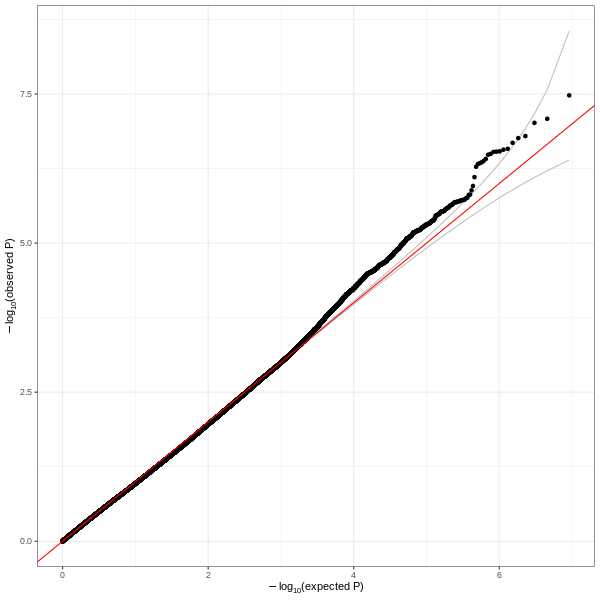

#### Primary studies: MVP

##### European-ancestry population

Supplementary Figure 9. Mirrored Manhattan plot showing genetic associations with short sleep duration (top) and long sleep duration (bottom) in European MVP participants

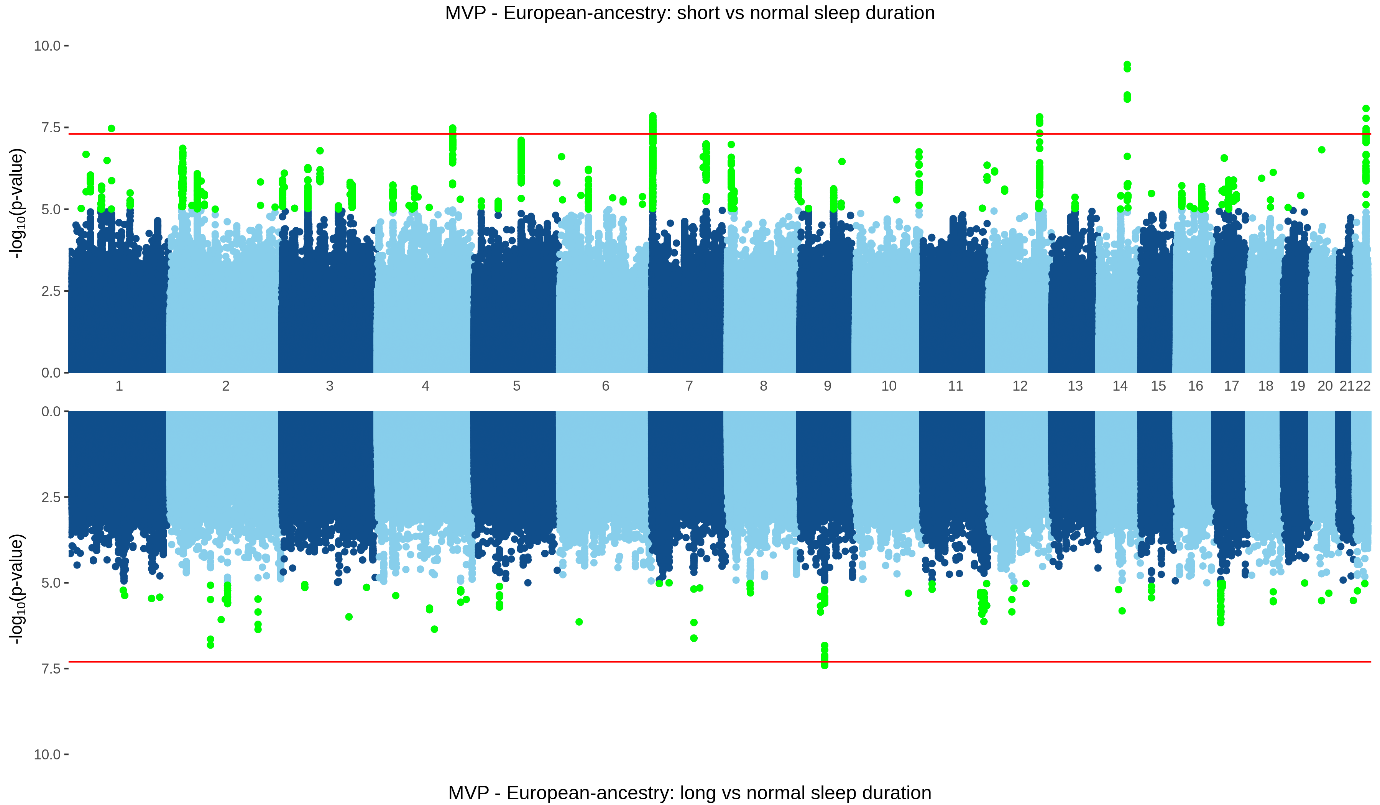

Supplementary Figure 10. Quantile-quantile plot for short sleep duration (left) and long sleep duration (right) in European MVP participants

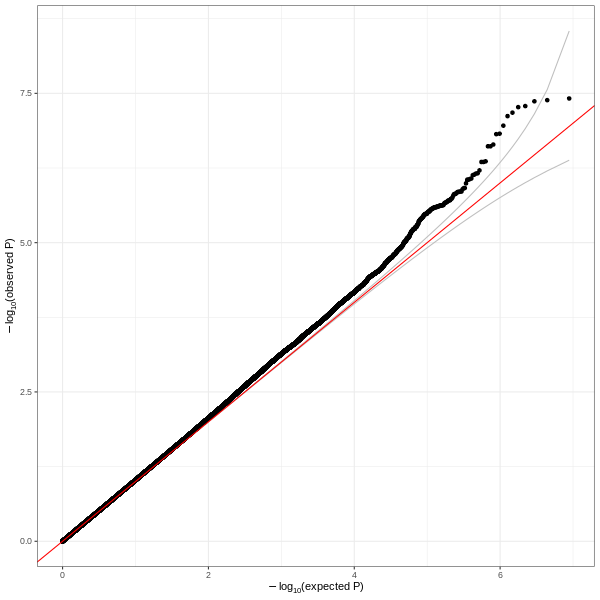

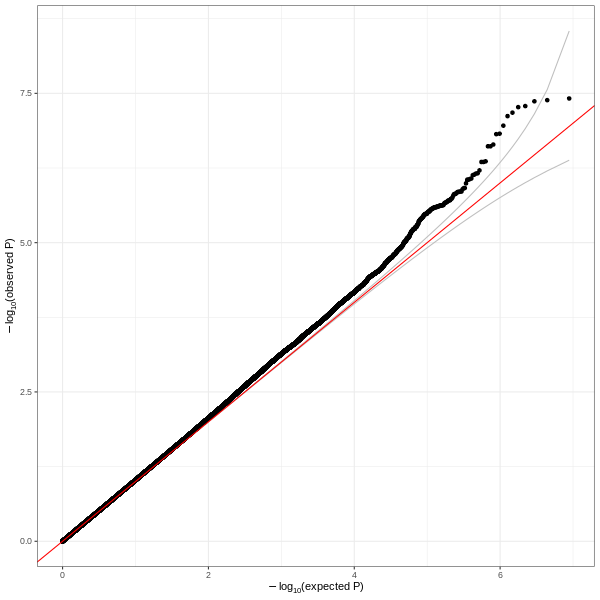

##### African-ancestry population

Supplementary Figure 11. Mirrored Manhattan plot showing genetic associations with short sleep duration (top) and long sleep duration (bottom) in African MVP participants

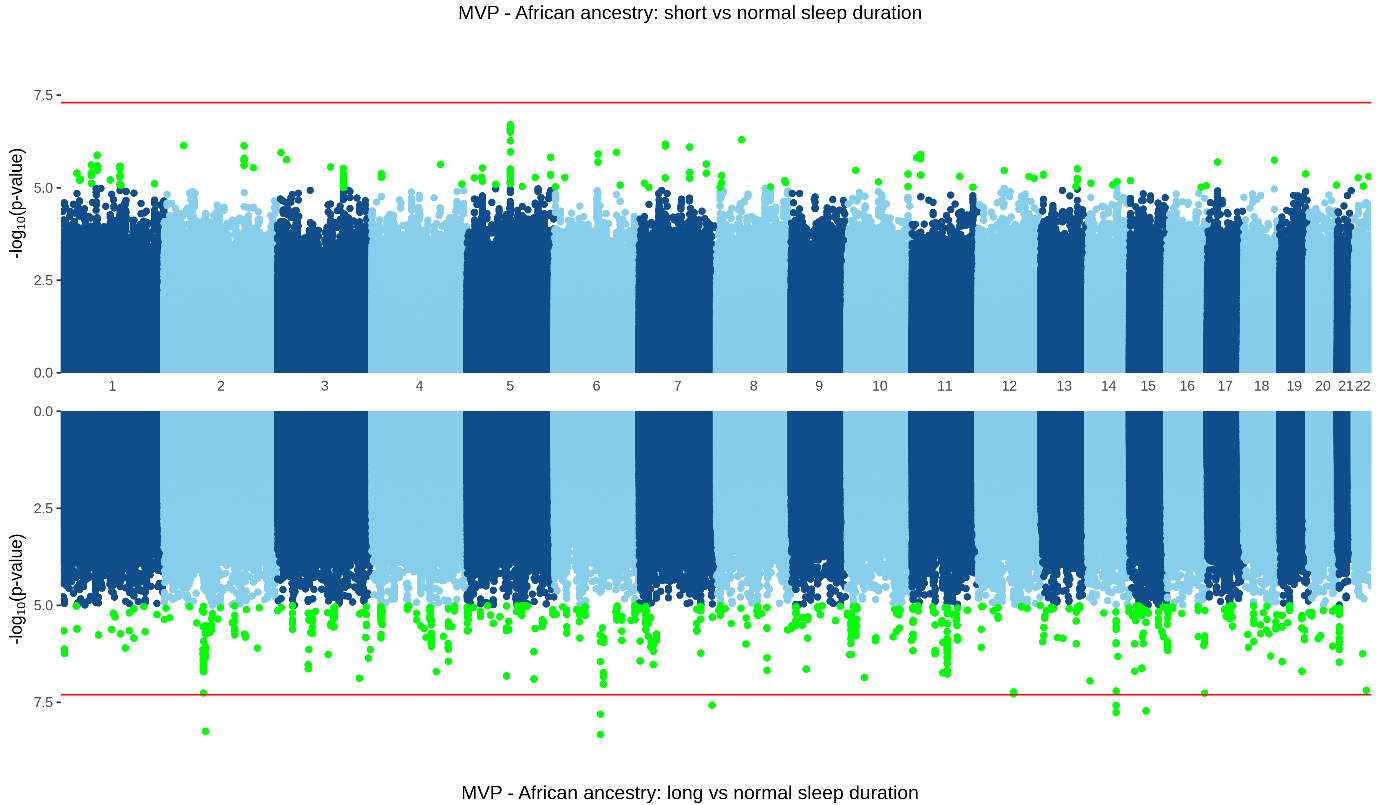

Supplementary Figure 12. Quantile-quantile plot for short sleep duration (left) and long sleep duration (right) in African MVP participants

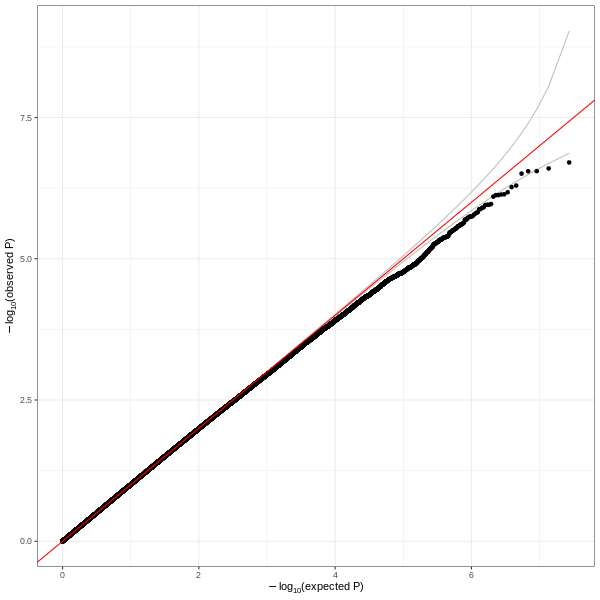

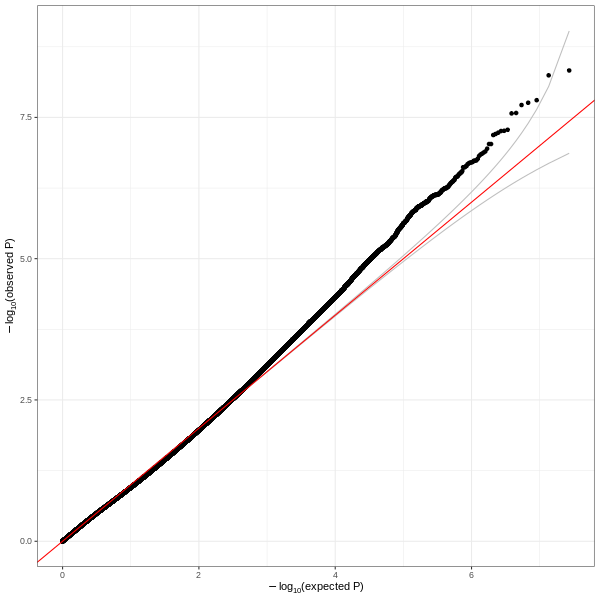

##### East Asian-ancestry population

Supplementary Figure 13. Mirrored Manhattan plot showing genetic associations with short sleep duration (top) and long sleep duration (bottom) in East Asian MVP participants

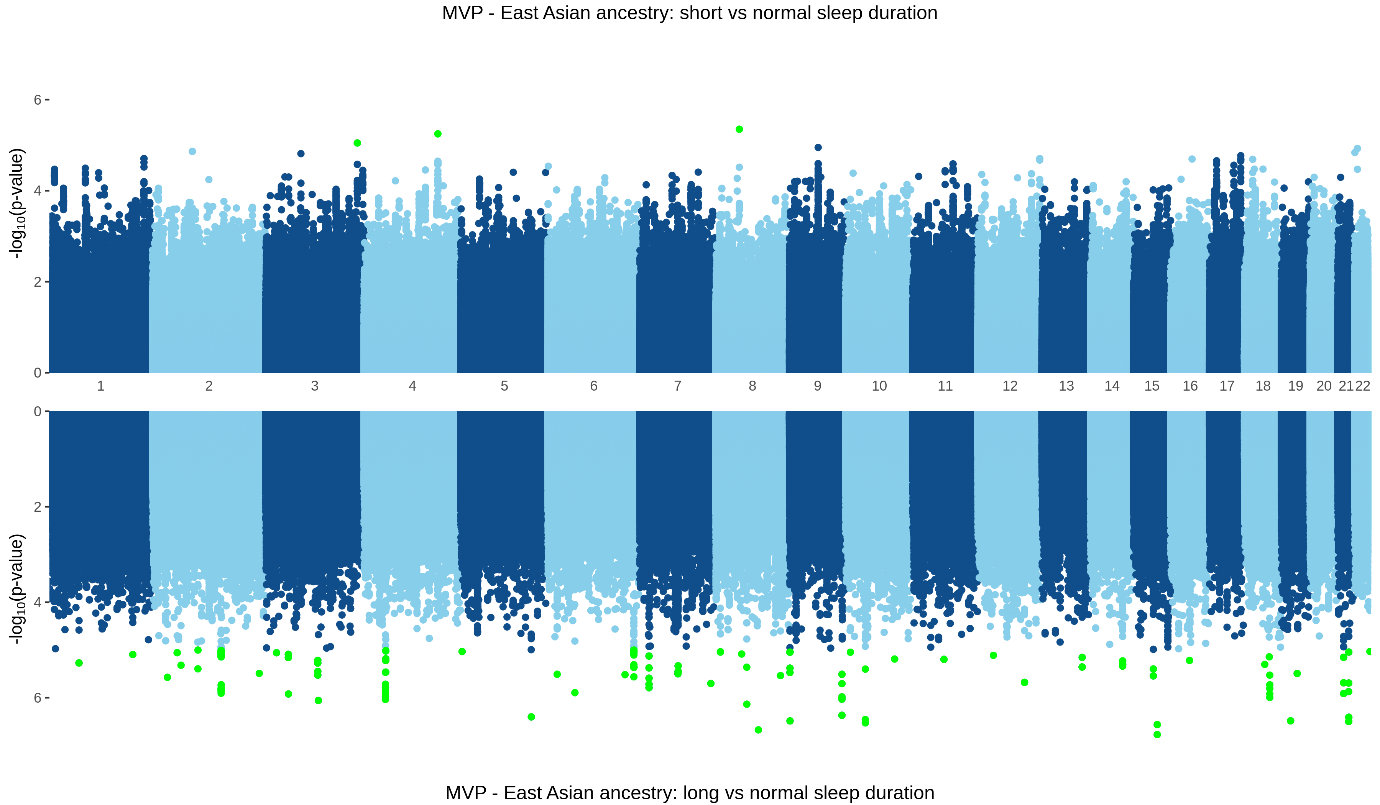

Supplementary Figure 14. Quantile-quantile plot for short sleep duration (left) and long sleep duration (right) in East Asian MVP participants

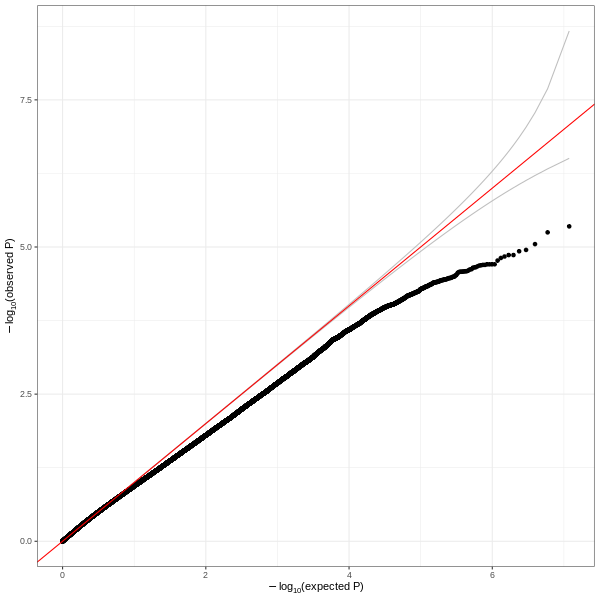

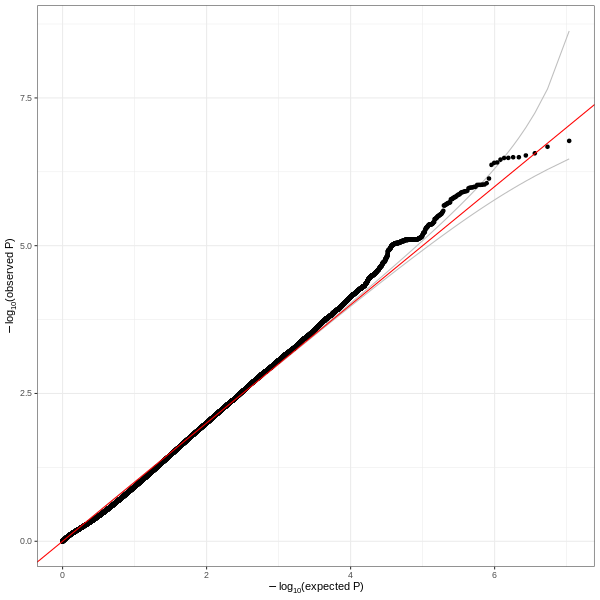

##### Admixed American population

Supplementary Figure 15. Mirrored Manhattan plot showing genetic associations with short sleep duration (top) and long sleep duration (bottom) in the East admixed-American MVP participants

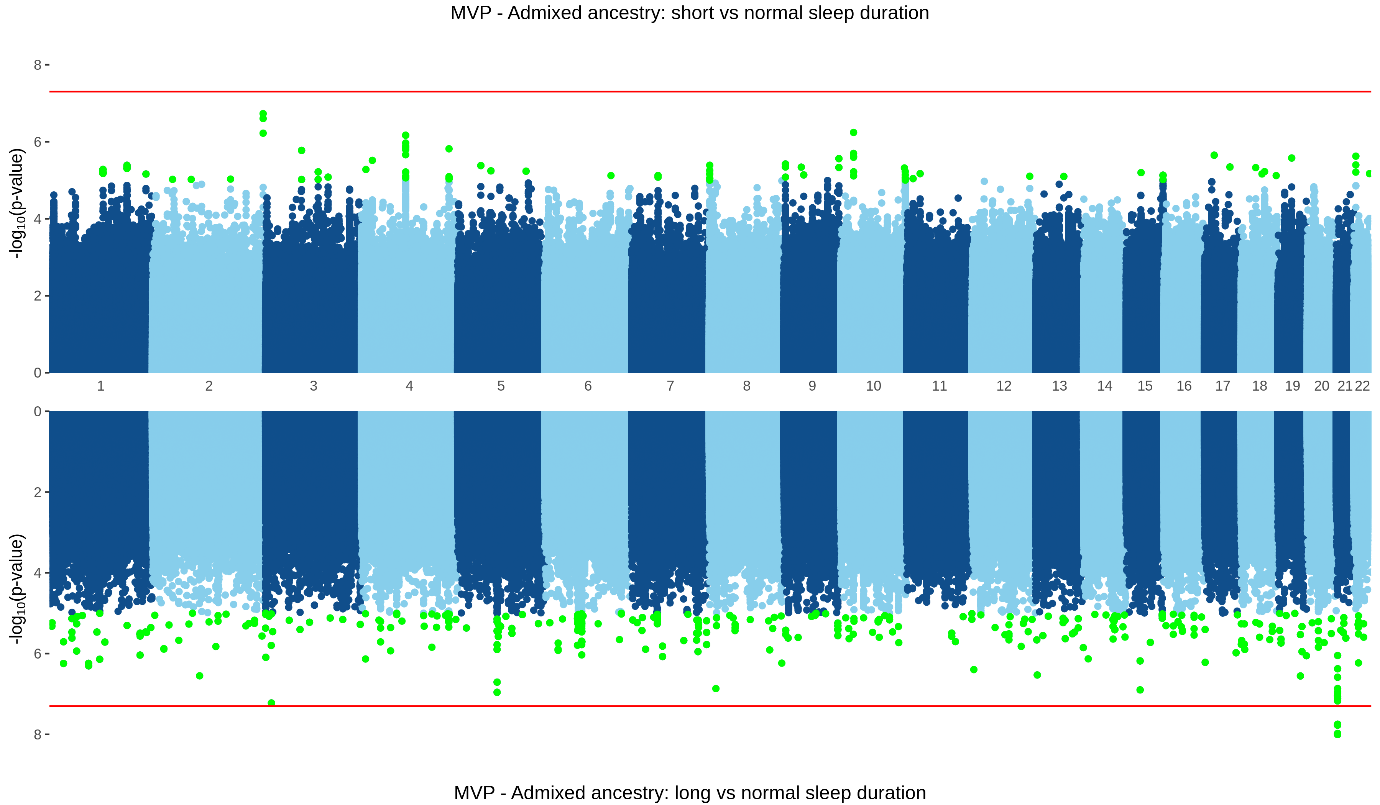

Supplementary Figure 16. Quantile-quantile plot for short sleep duration (left) and long sleep duration (right) in admixed-American MVP participants

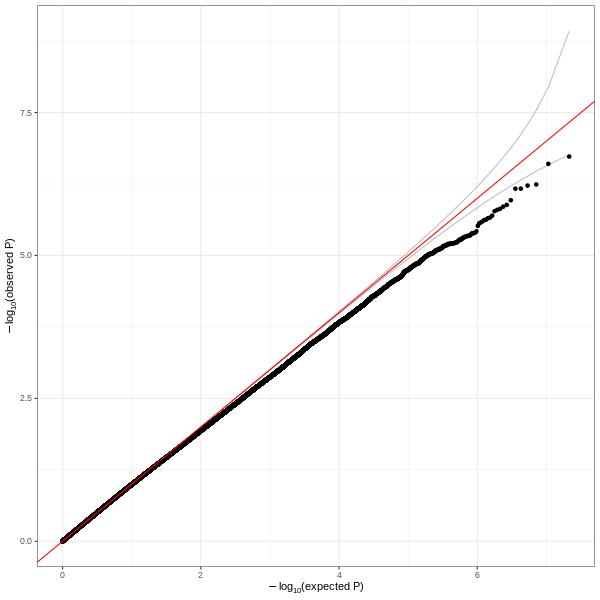

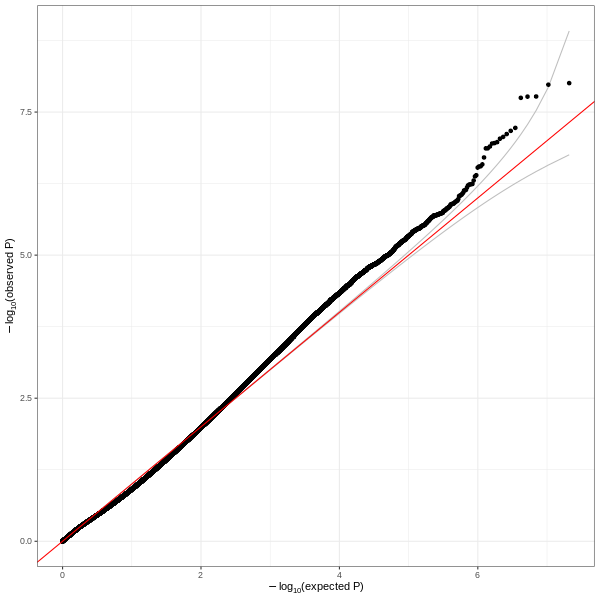

#### Ancestry-specific meta-analyses

##### European-ancestry population

Supplementary Figure 17 Mirrored Manhattan plot showing results of a meta-analysis in EUR groups in the UK Biobank and MVP cohorts. TOP: short (<6 hours, n=47,180) versus normal (7-8 hours, n=384,594), with 46 independent genetic-risk loci reaching genome-wide significance highlighted in green. BOTTOM: long (>9 hours, n=15,995) versus normal with one genome wide significant locus. All variants reaching a suggestive threshold of 1x10^-5^ are highlighted in green.

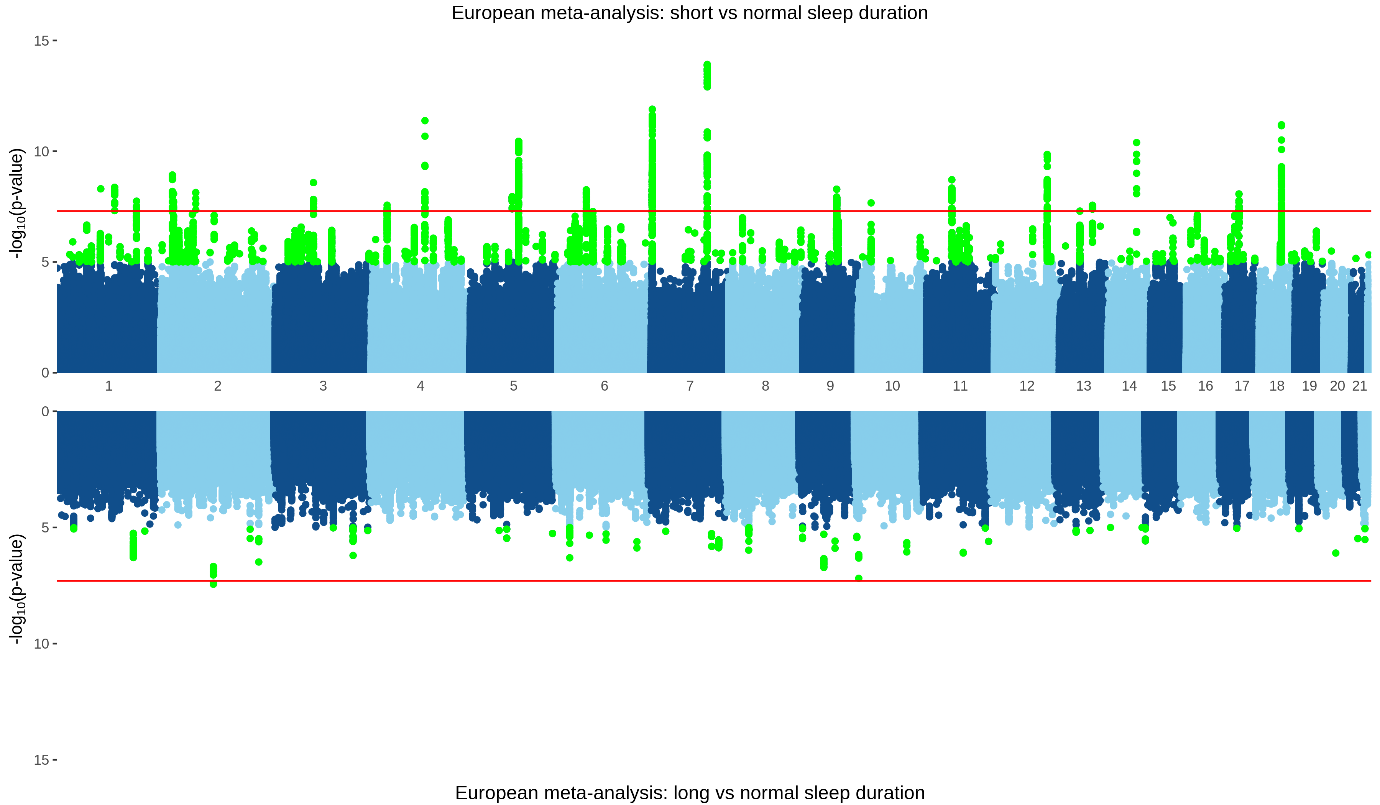

Supplementary Figure 18 Gene-based test for short (top) and long (bottom) sleep duration in EUR from UK Biobank and MVP cohorts. Bonferroni-adjusted significance threshold P<2.7x10^-6^

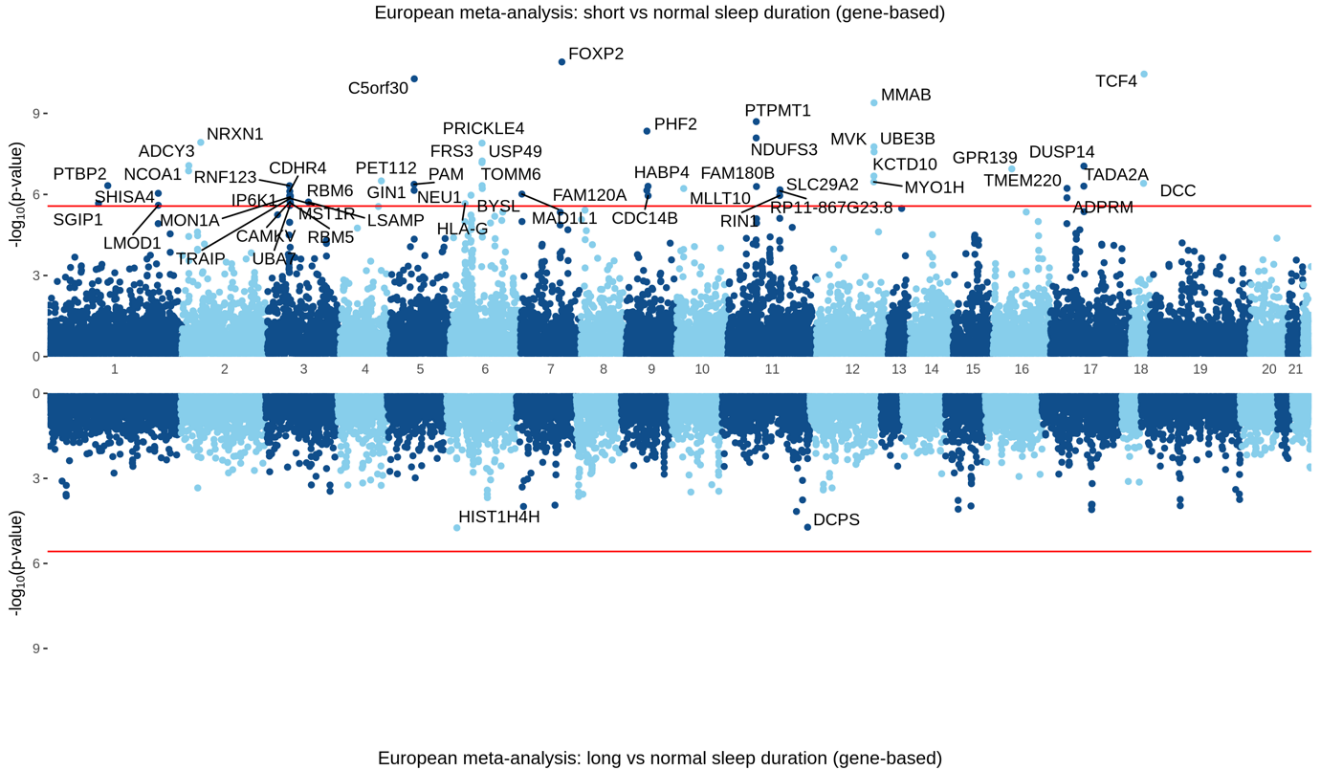

##### African-ancestry population

Supplementary Figure 19 Mirrored Manhattan plot showing results of a meta-analysis in AFR in the UK Biobank and MVP cohorts. TOP: short (<6 hours, n=11,352) versus normal (7-8 hours, n=15,305), with no genome-wide significance loci. BOTTOM: long (>9 hours, n=1,128) versus normal with one genome-wide significant locus. All variants reaching a suggestive threshold of 1x10^-5^ are highlighted in green.

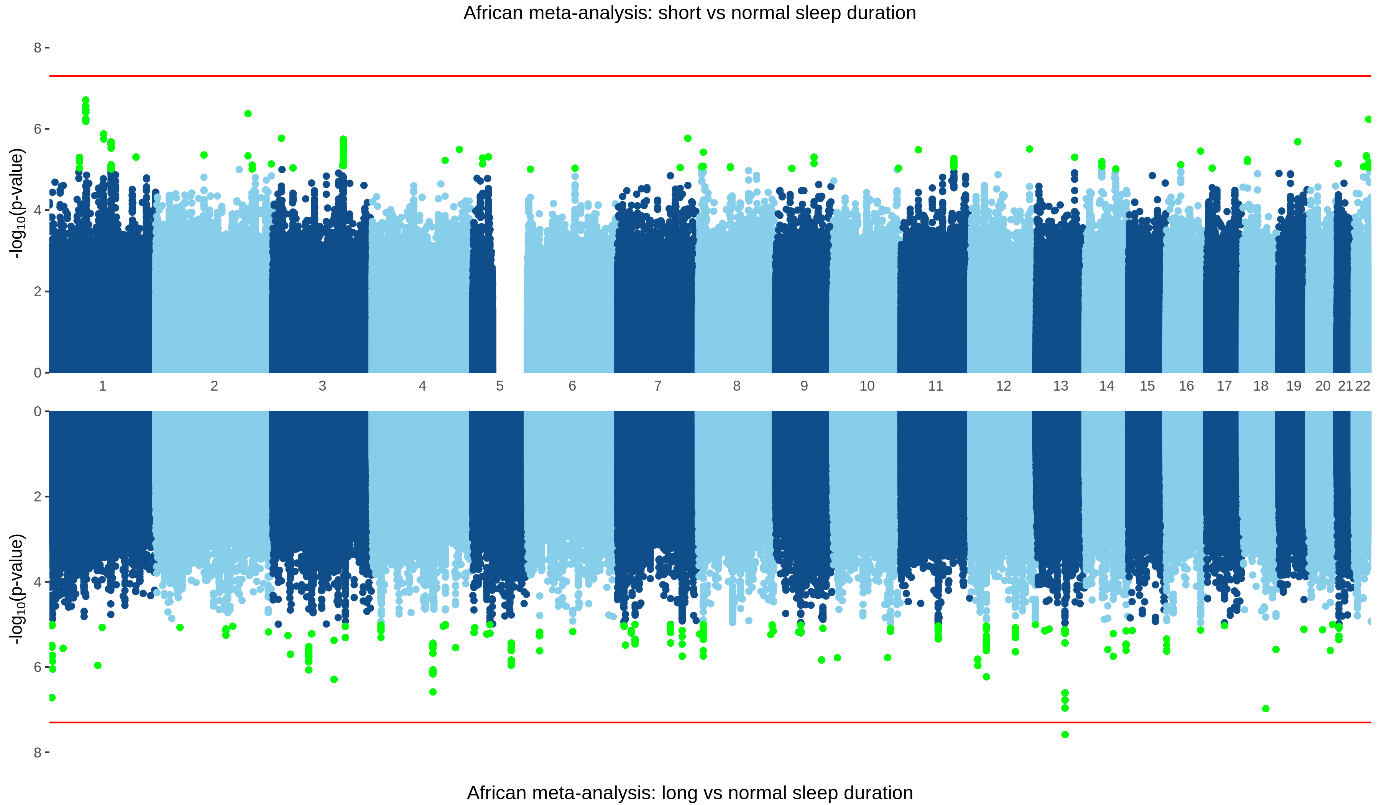

Supplementary Figure 20 Gene-based test for short (top) and long (bottom) sleep duration in African participants from UK Biobank and MVP cohorts, with top associations labelled.

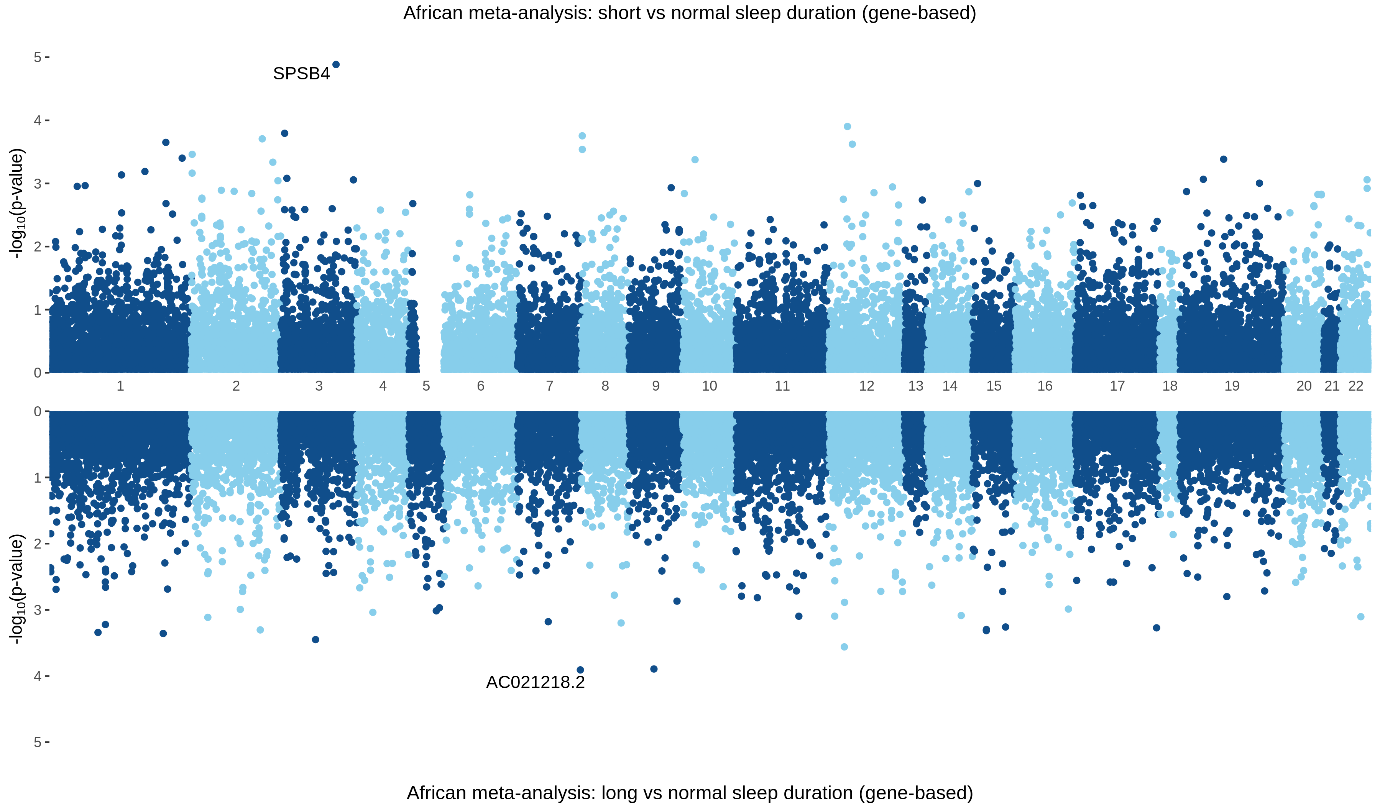

### Replication of previously published significant associations in the MVP sample

A previously published GWAS of sleep duration in European UK Biobank sample revealed several GWS risk loci for short sleep (<7 hours; 27 significant loci), long sleep (>8 hours; 8 significant loci) and sleep as a quantitative trait (from 0-24 hours) (78 significant loci) (14). Note that the definitions of short and long sleep differ slightly from our own. As noted in the main text, we did not assess the sleep data from MVP as a continuous trait. We used the European MVP-only GWAS to provide an independent replication sample for previously published GWAS assessing self-reported sleep duration as both a binary and quantitative trait in 446,118 European UK Biobank subjects^[1]^ (7). For this analysis, we considered a locus replicated if it reached a nominal significance threshold of p < 0.05 in the MVP GWAS. Where SNPs were not present in MVP, we investigated SNPs in high LD (r^2^>0.8) for replication, using 1000 Genomes Project European population as a reference panel. Where the primary study investigated sleep as a continuous trait, we considered a locus as replicated in our sample where the odds ratio was in the opposite direction for short sleep, and a consistent direction for long sleep

Of the 27 significant loci for short sleep duration, 17 were present in our study. We were able to identify reliable LD proxies for seven additional SNPs, leaving a total of three that could not be assessed in our data. A total of 10 reach a significance threshold of at least p < 0.05 in an independent sample of 158,222 EUR subjects from the MVP GWAS for short sleep duration (with a more stringent definition of <6 hours sleep) with same effect direction. In addition, two loci were significantly associated with long sleep in our MVP data, with an opposite direction of effect to that observed in the primary study of short sleep duration (see supplementary tables 17 and 18).

Of the eight loci significant associated with long sleep in (14), five are present in our MVP data, and we were able to identify a reliable LD proxy for a further two. One locus, rs549961083 on chromosome five, could not be assessed. None of these seven loci were replicated in our study of long sleep duration. However, two of these loci (rs4585442 on chromosome 5, and rs1229762 on chromosome 7) are significantly associated with short sleep duration in the MVP sample, with an opposite direction of effect to that reported in the original study of long sleep (see supplementary tables 19 and 20)

In addition to considering long and short sleep as binary traits, Dashti *et al* (14) conducted a GWAS of sleep as a quantitative continuous measure, for which they identify 78 GWS loci. A total of 58 of these loci were present in our MVP data, and we identified reliable LD proxies for further 12. Eight of these loci could not be assessed in our data. Of those 78 significant associations with continuous sleep duration, we replicate a total of 18 of these associations in our MVP GWAS on short sleep, and three in our MVP GWAS on long sleep (see supplementary tables 21 and 22).

1. Dashti HS, Jones SE, Wood AR, Lane JM, van Hees VT, Wang H, et al. Genome-wide association study identifies genetic loci for self-reported habitual sleep duration supported by accelerometer-derived estimates. Nat Commun. 2019;10(1):1100.
